# Joint Bayesian modelling of molecular QTL and GWAS effects improves polygenic prediction for complex traits

**DOI:** 10.64898/2026.03.10.26347908

**Authors:** Shouye Liu, Yang Wu, Zhili Zheng, Hao Cheng, Michael E. Goddard, Jian Yang, Peter M. Visscher, Jian Zeng

**Affiliations:** Institute for Molecular Bioscience, The University of Queensland, Brisbane, Queensland, Australia; Institute of Rare Diseases, West China Hospital of Sichuan University, Chengdu, China; Program in Medical and Population Genetics, Broad Institute of Harvard and MIT, Cambridge, Massachusetts, USA; Stanley Center for Psychiatric Research, Broad Institute of Harvard and MIT, Cambridge, Massachusetts, USA; Analytic and Translational Genetics Unit, Massachusetts General Hospital, Boston, Massachusetts, USA; Faculty of Veterinary and Agricultural Science, University of Melbourne, Parkville, Victoria, Australia; Biosciences Research Division, Department of Economic Development, Jobs, Transport and Resources, Bundoora, Victoria, Australia; Department of Animal Science, University of California Davis, Davis, CA95616, USA; New Cornerstone Science Laboratory, School of Life Sciences, Westlake University, Hangzhou, Zhejiang, China; Westlake Laboratory of Life Sciences and Biomedicine, Hangzhou, Zhejiang, China; Nuffield Department of Population Health, University of Oxford, Oxford, United Kingdom

**Author notes:** Correspondence: Shouye Liu < >, Jian Zeng < >.

## Abstract

Integrating molecular quantitative trait locus (molQTL) data into polygenic prediction offers a promising strategy for improving complex trait prediction. We introduce SBayesCO, a Bayesian framework that jointly models genome-wide association study (GWAS) and molQTL effect sizes by treating the complex trait and molecular phenotypes as genetically correlated traits. SBayesCO estimates genome-wide SNP effects on both phenotypes and supports analyses using individual- or summary-level data. In simulations, SBayesCO consistently outperforms SBayesC, a baseline model based solely on GWAS data, especially when GWAS sample sizes are modest. Applying to 11 blood and immune-related traits using large-scale expression QTLs (eQTLs) and protein QTLs (pQTLs) resources, SBayesCO achieves relative improvements in prediction R^2^ of up to 6.3% with pQTLs and 5.3% with eQTLs compared to SBayesC, with similar gains relative to modelling molQTLs as binary annotations (SBayesCC). SBayesCO also improves SNP prioritization by concentrating posterior inclusion probabilities on regulatory variants. These results demonstrate the value of modelling quantitative molQTL effect sizes and provide guidance on how increasingly available functional genomic annotations, including AI-based regulatory effect predictions, can be effectively integrated to improve polygenic prediction.

## Introduction

The genetic architecture of complex traits is shaped by numerous variants of modest effects acting across diverse biological pathways and regulatory layers^1,2^. Although GWAS have identified thousands of trait-associated variants^3^, accurate polygenic prediction remains challenging because variants reaching genome-wide significance are often not causal and collectively explain only a limited proportion of SNP-based heritability^4,5^. Methodological advances in random effects models, e.g., LDpred^6^, BayesR^7^, SBayesR^8^, PRS-CS^9^, and others^10-16^, have improved prediction accuracy by jointly fitting all genome-wide SNPs while accounting for linkage disequilibrium (LD). These models typically assume a sparse genetic architecture through mixture or shrinkage priors on SNP effects. However, they generally do not incorporate gene regulatory information, which may be critical for distinguishing causal variants from tagging SNPs and for characterizing the underlying causal effect distribution, thereby improving polygenic prediction of complex traits^17^.

Recent efforts have extended polygenic prediction models by incorporating functional annotations^18^, including AnnoPred^19^, PolyFun^20^, LDpred-funct^21^, BayesRC^22^, and SBayesRC^23^. These methods represent an important advance by modelling annotation-informed effect size variances or causal probabilities, enabling probabilistic and context-aware SNP prioritisation^21,24^. However, the functional annotations used are often generic, such as promoter or enhancer sequences, and are not specific to molecular phenotypes such as gene expression or protein abundance^25^. Moreover, most annotation-informed models treat annotations as a categorical indicator of relevance rather than modelling the quantitative contribution of regulatory mechanisms^26^. This simplification limits their ability to distinguish between SNPs with strong versus weak regulatory effects.

Unlike general annotations such as sequence conservation^27^ or chromatin marks^28^, molQTLs provide direct measurements of how genetic variation influences molecular phenotypes, thereby offering a mechanistic link between non-coding genetic variation and downstream functional consequences^29^. Importantly, molQTLs provide not only evidence of regulatory activity but also effect size estimates that quantify the magnitude and direction of regulatory influence^30,31^. Recent fine-mapping studies have shown that incorporating molQTL effect sizes, rather than their mere presence or absence, substantially improves power and resolution for identifying likely causal variants^32,33^. Although gene-based scoring approaches like TWAS can be used to improve prediction^34-36^, these methods typically aggregate signals at the gene level, ignore intergenic variants, and are not designed for genome-wide SNP-level modelling. Consequently, despite their interpretability and mechanistic relevance, molQTL effect sizes remain underutilised in current polygenic prediction models.

In this study, we introduce SBayesCO, a joint Bayesian mixture model that improves polygenic prediction by explicitly integrating molQTL and GWAS effects at the genome-wide SNP level. The model treats a complex trait and relevant molecular phenotypes (e.g., gene expression or protein abundance) as genetically correlated traits, enabling joint inference of trait-shared and trait-specific effects within genic regions. To capture additional genetic associatioin signals, intergenic SNPs are modelled as joint components. SBayesCO is implemented in our recently developed BayesOmics suite, a user-friendly, versatile C++ software supporting both individual-level and summary-level analyses, LD reference construction, and molQTL data management (https://shouyeliu.github.io/softwares/content-softwares.html). We evaluated the performance of SBayesCO using simulations with different scenarios and applications to 11 UK Biobank (UKB) traits, leveraging pQTLs from the UKB Pharma Proteomics Project (UKB-PPP) project^37^ and eQTLs from the eQTLGen consortium^38^. We systematically compared three modelling strategies: i) SBayesC^39^, a baseline model without annotations; ii) SBayesCC, a variant of SBayesRC that incorporates binary annotations indicating whether a SNP lies within the cis region (±1 Mb) of a molecular phenotype via a point-normal prior; and iii) our proposed model, which uses the same cis-region molQTL data but directly leverages quantitative molQTL effect sizes and a bivariate prior to model SNP inclusion, effect variance, and trait-molecular covariance. These analyses aim to inform how quantitative molQTL and other functional genomic information can be most effectively incorporated into polygenic prediction of complex traits.

## Results

### SBayesCO model overview

SBayesCO is a Bayesian bivariate mixture model that incorporates quantitative molQTL information (molQTL effect sizes and their standard errors) from the cis-region (± 1Mb), while jointly modelling all genome-wide SNPs (Fig. 1b). The model aims to improve polygenic score prediction by leveraging molQTL effects to identify variants influencing both complex trait and molecular phenotypes, accounting for both trait-shared and trait-specific genetic effects within genic regions, alongside residual polygenic effects from intergenic SNPs.

**Fig. 1.**
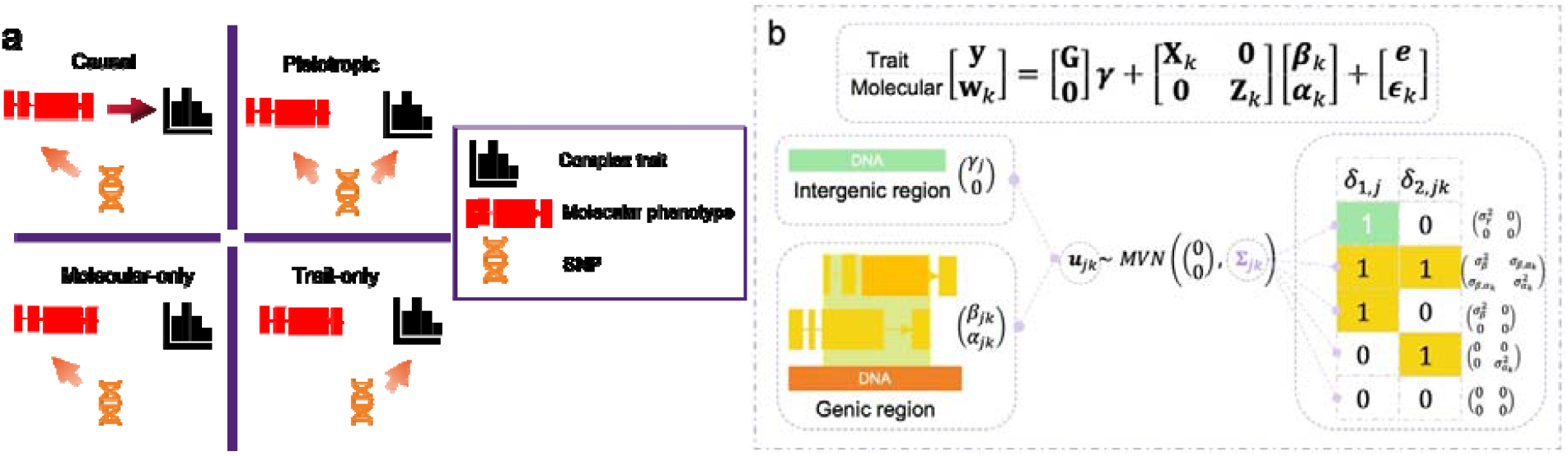
Schematic overview of SBayesCO. a) Four possible relationships among genotype, a molecular phenotype, and a complex trait. Top left (causal mediation): the SNP influences the complex trait directly through regulation of the molecular phenotype. Top right (pleiotropy): the SNP has direct effects on both the molecular phenotype and the complex trait. Bottom left (molecular-only): the SNP affects only the molecular phenotype. Bottom right (trait-only): the SNP affects only the complex trait. b) Model structure and assumptions underlying SBayesCO-EIEIO. SBayesCO is a unified Bayesian framework that integrates GWAS and molQTL data to improve polygenic prediction. To accommodate heterogeneity across SNPs, the genome is partitioned into genic and intergenic regions, with region-specific spike-and-slab priors. Within genic regions, each cis-SNP may influence the complex trait, one or more molecular phenotypes, or both. SNPs in intergenic regions are modelled as a special case of the bivariate framework with molQTL effects fixed to zero ( ). Overall, SBayesCO jointly models correlated and uncorrelated genetic effects between the complex trait and molecular phenotypes using a bivariate normal mixture prior.

In contrast to SBayesCC (two-component SBayesRC^23^), which incorporates binary or quantitative annotation to weigh single-trait effects, SBayesCO directly integrates quantitative functional information through a bivariate framework. The genome is partitioned into genic and intergenic regions, with region-specific priors reflecting distinct biological roles. Within genic regions, each cis-SNP is allowed to affect the complex trait, the molecular phenotype, or both, capturing both pleiotropic and molecularly mediated effects. SNPs located in intergenic regions are modelled jointly, without direct molecular effects, to capture remaining polygenic signals not explained by molQTL-integrated components.

### Assessing SBayesCO performance through simulations

We assessed the performance of SBayesCO using simulated quantitative phenotypes for both the complex trait and gene expression, based on HapMap3^40^ SNPs on chromosome 22 and gene positional information (Methods; Fig. 1a). Under the causal mediation scenario, where SNP effects on the complex trait operated through regulation of gene expression, individual-level models (BayesCO and BayesC) consistently outperformed their summary-level counterparts across all methods (Fig. 2a), as individual-level data retained the full amount of genetic information.

**Fig. 2.**
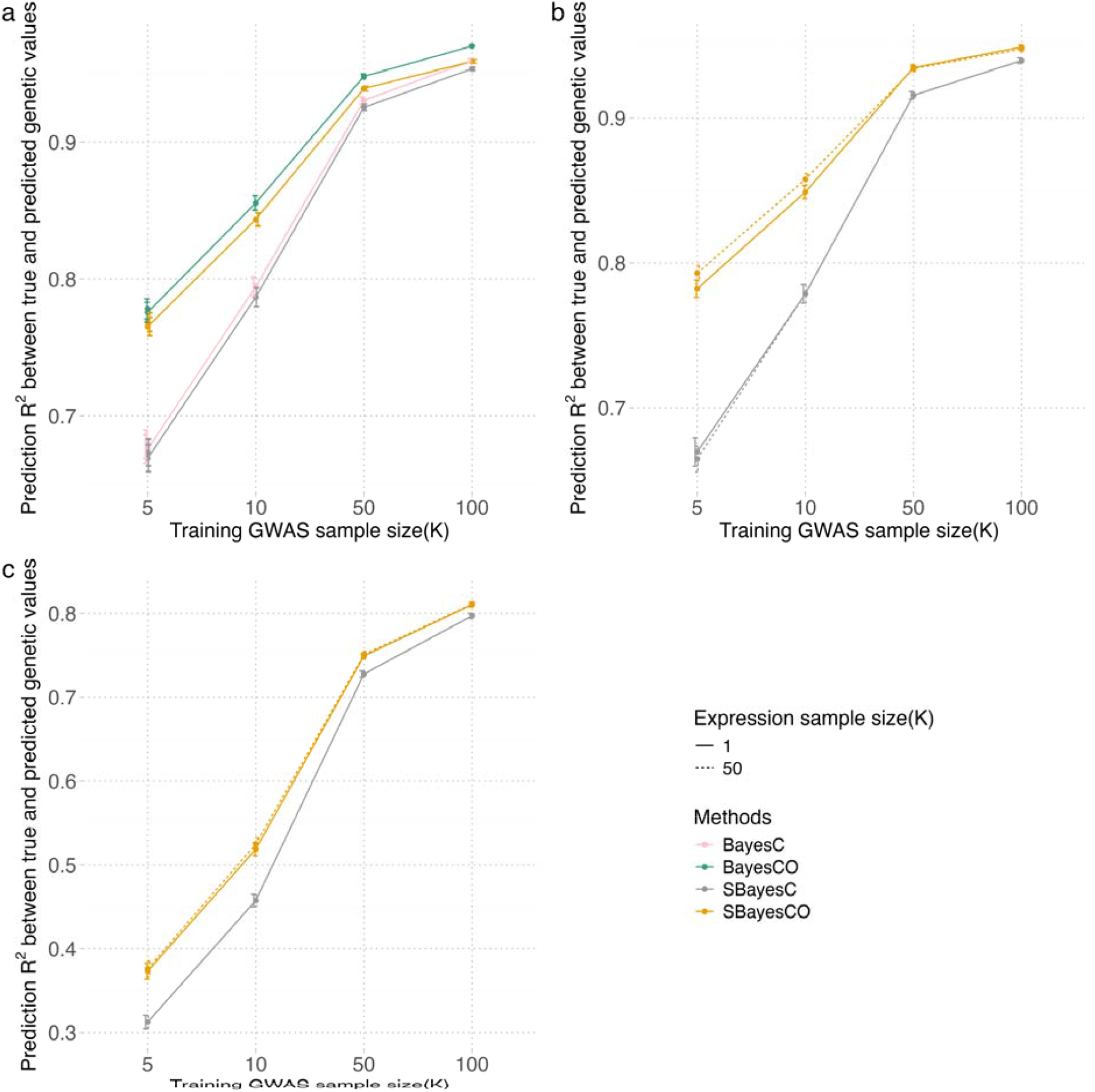
Prediction accuracy in simulation studies. Simulation studies were conducted under two main scenarios that varied in the number of causal variants and genes, as well as in sample sizes for the complex trait and gene expression (Methods). Prediction accuracy was evaluated by prediction *R*^2^ between true and predicted genetic values. a) Comparison of individual-level and summary-level methods using internal LD and eQTL data in the causal scenario. b) Prediction performance in the causal scenario using external LD and eQTL datas. c) Prediction performance in the realistic scenario using external LD and eQTL data.

Across all sample-size settings, SBayesCO outperformed the baseline SBayesC model, with increasing differences as GWAS sample size decreased (Fig. 2a). This pattern highlights the benefit of integrating orthogonal molQTL information, particularly for underpowered GWAS. Boosting the eQTL sample size from 1K to 50K further improved prediction accuracy, with the largest gains observed when GWAS sample size was small. For instance, by incorporating eQTL data from 50K samples, SBayesCO trained on a GWAS of 5K individuals achieved higher prediction *R*^2^ than SBayesC trained on a GWAS of 10K individuals (Fig. 2b).

Under a more realistic genetic architecture (Fig. 2c), where causal variants can influence gene expression and the complex trait, separately, or influence both via mediation or pleiotropy (Methods), SBayesCO consistently achieved higher predictive accuracy than SBayesC across a range of GWAS sample sizes. However, increasing the eQTL sample size did not lead to additional improvement in prediction accuracy. These results can be understood as follows. Under causal mediation, SNP effects on the trait are partially determined by their effects on gene expression, so increasing the eQTL sample size improves the estimation of eQTL effects and gene-trait covariance, thereby increasing prediction accuracy. Under pleiotropy, by contrast, SNP effects on expression and on the trait are independent, so even highly precise eQTL estimates provide little information about trait effects. Consequently, while eQTL data help prioritize regulatory variants, further increases in eQTL sample size have little additional impact on trait prediction.

### Modelling eQTL effects improves prediction over binary annotation

To evaluate the performance of SBayesCO using real data, we conducted five-fold cross-validation across 11 independent blood and immune-related traits, using approximately 1 million HapMap3 SNPs and unrelated individuals of European (EUR) ancestry (average sample size N=327,569 across traits). We used GWAS summary statistics from the UKB, together with cis-eQTL data from the eQTLGen consortium^38^ (72.3% of the SNPs in cis; Methods; Supplementary Tables 1–2). Predictive performance was benchmarked against two Bayesian models: SBayesC and SBayesCC^23^ (Fig. 3; Supplementary Table 3-4). To facilitate comparisons across traits with different genetic architectures, prediction accuracy was standardized by dividing prediction *R*^2^ by SNP-based heritability 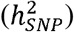, and relative improvement was computed with respect to SBayesC.

**Fig. 3.**
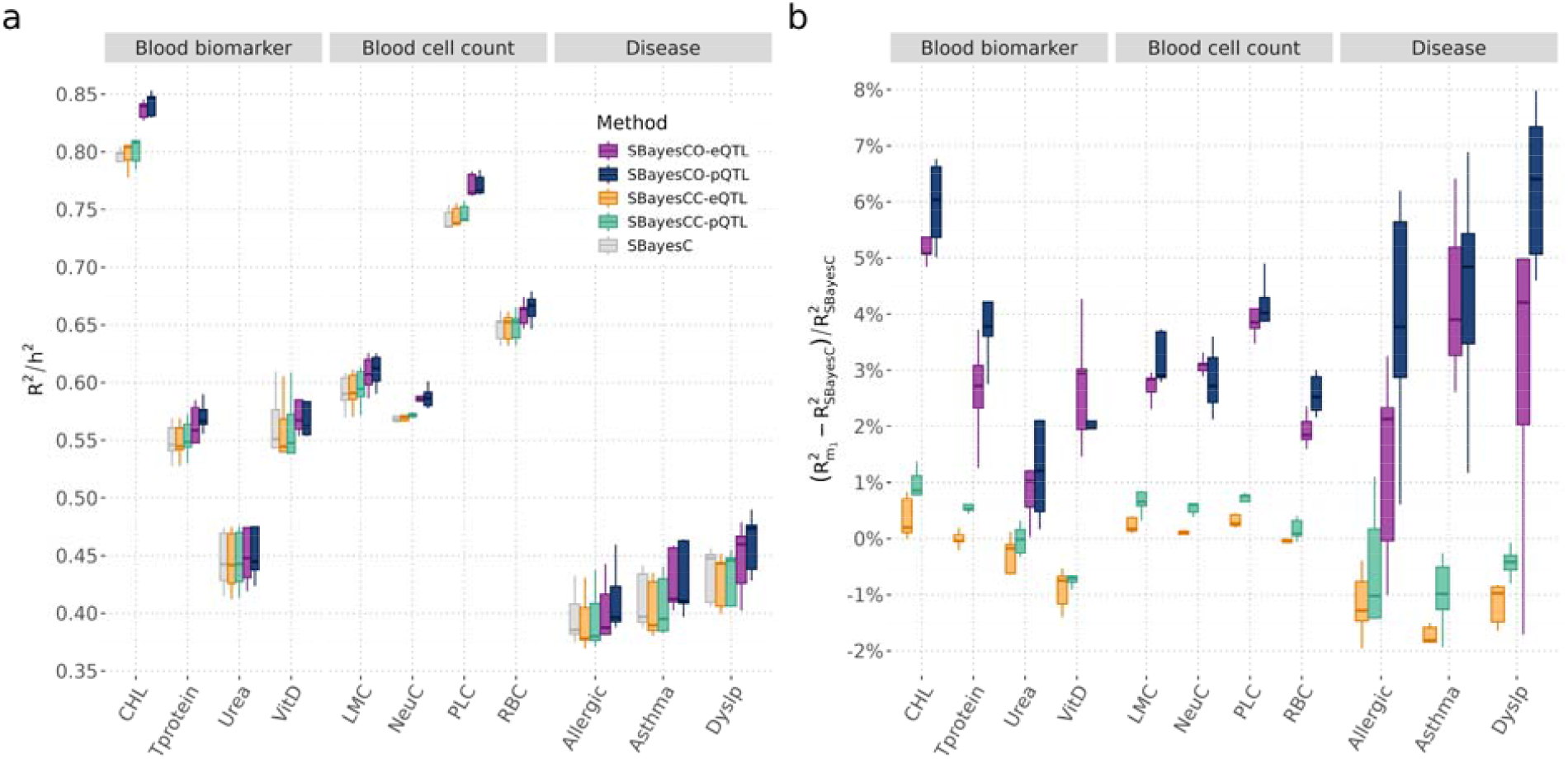
Prediction accuracy across 11 UKB blood-related traits with molQTL data in blood. a) Standardised prediction accuracy 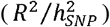, where *R*^2^ is the out-of-sample prediction accuracy and 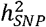 is the SNP-based heritability estimated using SBayesC. b) Relative gain in *R*^2^ compared with SBayesC, calculated as 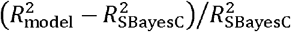, where model ∈ [ SBayesCC, SBayesCO]. This metric quantifies the proportional improvement in prediction accuracy. Positive values indicate improved performance relative to SBayesC. Different fill colours denote prediction methods.

SBayesCO consistently outperformed both SBayesC and SBayesCC across all 11 traits. Averaged across traits, SBayesCO increased 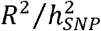 by 2.9% relative to SBayesC (range: 0.8–5.3%) and by 3.3% relative to SBayesCC (range: 1.1–6.1%). Improvements in absolute prediction accuracy were also observed, with mean relative gains in R^2^ of 2.8% for diseases and 2.9% for both blood biomarkers and blood cell count traits (Fig. 3b). In contrast, SBayesCC yielded only modest benefits relative to SBayesC, with a mean increase in 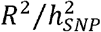 of only 0.2% for blood cell traits and an average decrease for diseases (–1.4%), suggesting limited effectiveness of binary annotation modelling for complex genetic architectures.

Examples from individual traits further illustrate the consistent advantages of SBayesCO. For total cholesterol (CHL), a blood biomarker, SBayesCO increased 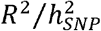 from 0.794 under SBayesC to 0.837 (a 5.3% relative increase), whereas SBayesCC produced only a marginal gain (0.797; 0.37%). For asthma, a complex disease, SBayesCO improved 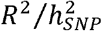 from 0.410 to 0.428 (4.3%), whereas SBayesCC slightly reduced the standardized prediction accuracy (0.403; –1.7%). Similarly, for platelet count (PLC), a blood cell trait, SBayesCO increased 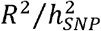 from 0.741 in SBayesC to 0.771 (4.0%), compared to a minimal improvement under SBayesCC (0.744; 0.3%).

Together, these results demonstrate that although incorporating eQTL information can improve polygenic prediction, the magnitude and robustness of the improvement critically depend on how molecular information is modelled. Treating eQTL information as binary annotation yielded limited gains and sometimes reduced prediction accuracy, because a single annotation inevitably mixes SNPs that influence both gene expression and the trait with those that only regulate expression. This mixture can inflate false positives and dilute trait-relevant signals. In contrast, SBayesCO directly models quantitative eQTL effect sizes, allowing the trait-gene genetic covariance to be estimated and used to weight the contribution of eQTL effects to trait-effect estimation. This leads to larger and more consistent improvements in prediction performance. We next assess whether this strategy generalises to other molecular layers by incorporating pQTL effect sizes.

### Incorporating pQTL data leads to greater improvements across traits

We assessed prediction accuracy across the same 11 traits using cis-pQTL data from the UKB-PPP project^37^ (69.0% of the SNPs in cis regions). Incorporating pQTL effect sizes within the SBayesCO framework consistently improved prediction accuracy over both SBayesC and SBayesCC (Fig. 3; Supplementary Table 3-4). Averaged across traits, SBayesCO increased 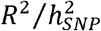 by 3.7% compared to SBayesC (range: 1.2–6.3%) and by 3.6% compared to SBayesCC (range: 1.2–6.7%), slightly higher than those observed using eQTL information. When evaluated using prediction accuracy, SBayesCO-pQTL achieved a mean relative improvement in *R*^2^ of 3.7% over SBayesC, exceeding the gain obtained with SBayesCC-pQTL (0.08%). Stratified by trait category, the mean relative improvement under SBayesCO-pQTL, relative to SBayesC, was 4.8% for diseases, 3.4% for blood biomarkers, and 3.2% for blood cell count traits.

Although the overall differences in prediction accuracy between using eQTL and pQTL data were modest, pQTL yielded higher accuracy for most traits across both SBayesCC and SBayesCO (Fig. 3), suggesting that pQTL may provide more trait-relevant regulatory information, potentially because protein abundance lies closer to complex trait biology than gene expression. Consistent with this, SBayesCO-pQTL showed robust improvements when quantitative effect sizes were incorporated, whereas SBayesCC-pQTL using binary annotation showed only limited improvements. For example, across disease traits, the average relative improvement in *R*^2^ over SBayesC was 4.8% for SBayesCO-pQTL and 2.8% for SBayesCO-eQTL, whereas the corresponding SBayesCC models reduced prediction *R*^2^ on average (–1.35% for SBayesCC-eQTL and –0.64% for SBayesCC-pQTL).

Trait-level comparisons further illustrate the consistent advantages of SBayesCO that models effect sizes through a bivariate framework. The largest improvement using pQTL information was observed for dyslipidemia (Dyslip; 6.3%), whereas the largest improvement using eQTL information was observed for total cholesterol (5.3%). Together, these results indicate that modelling quantitative molQTL effects yields more robust and reproducible improvements in polygenic prediction than treating molecular information as binary annotations.

### Improved prediction portability in South Asian ancestry

We conducted trans-ancestry prediction analyses to assess the portability and robustness of molQTL–integrated polygenic models beyond EUR ancestry (Fig. 4; Supplementary Table 5). In these analyses, SNP effect sizes were estimated using the full set of European individuals as the training data, and prediction was evaluated in independent non-European target populations, including African (AFR; *n* = 7,015), East Asian (EAS; *n* = 2,256), and South Asian (SAS; *n* = 9,448). When prediction was evaluated on EUR validation samples, incorporating molQTL effect sizes yielded relatively consistent gains across traits, although the overall magnitude of improvement was modest (Fig. 3). In contrast, applying the same EUR-trained effects to non-European populations resulted in substantially greater heterogeneity and ancestry-dependent variation in prediction accuracy, with some traits showing improvement of up to 38% in EAS and others showing minimal gains or even reduced accuracy. These patterns likely reflect differences in genetic distance to EUR ancestry and the reliance on common HapMap3 SNPs, which limits the ability to capture causal variants across populations. When causal variants are not observed, larger differences in allele frequencies and linkage disequilibrium structure across populations reduce trans-ancestry portability and increases instability in prediction performance.

**Fig. 4.**
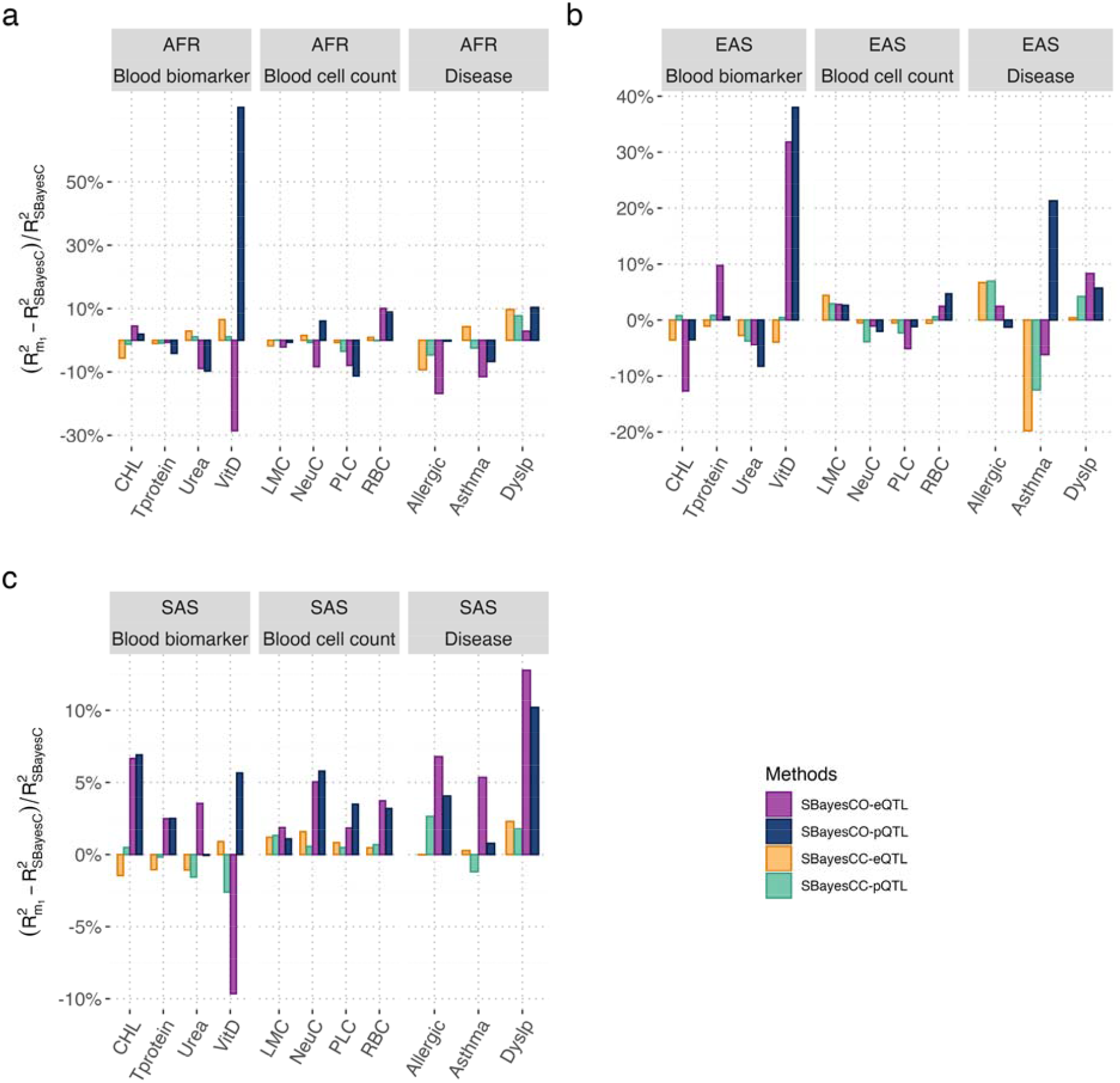
Cross-ancestry prediction accuracy relative to baseline (SBayesC) model. Relative changes in prediction accuracy are shown for African (AFR; a), East Asian (EAS; b), and South Asian (SAS; c) target populations. Traits are grouped into blood biomarkers (CHL, Tprotein, Urea, VitD), blood cell counts (LMC, NeuC, PLC, RBC), and immune-related diseases (Allergic, Asthma, Dyslip) (Supplementary Table 1). Bars represent the relative gain in prediction accuracy compared with the baseline SBayesC model, calculated as, calculated as 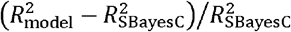, where model ∈ [ SBayesCC, SBayesCO]. This metric quantifies the proportional improvement in prediction accuracy. Positive values indicate improved performance relative to SBayesC. Different fill colours denote prediction methods.

In both AFR and EAS populations, cross-ancestry polygenic prediction using EUR-trained SNP effect sizes showed little evidence of systematic improvement over the annotation-free baseline (SBayesC). Across blood biomarkers, blood cell count traits, and disease phenotypes, relative changes in prediction accuracy were generally small, inconsistent in direction, and often close to zero or negative. Vitamin D (VitD) exhibited pronounced relative changes under certain molQTL– integrated models; however, these fluctuations arose from extremely low baseline prediction accuracy in these populations. Because AFR and EAS are genetically more distant from EUR, relative differences become highly sensitive to small absolute changes.

In contrast, prediction results in the SAS population, which is genetically closest to EUR among the non-EUR considered, were notably more consistent across traits and modelling strategies. SBayesCC-eQTL and SBayesCC-pQTL exhibited notable trait-category–dependent behaviour. Consistent with results within EUR, these models generally improved prediction relative to SBayesC for blood cell count traits, but had limited or even negative effects for blood biomarker traits and disease phenotypes, suggesting that annotation-based modelling can yield heterogeneous and sometimes adverse effects across trait categories. By comparison, SBayesCO models that directly incorporate eQTL or pQTL effect sizes produced more robust and broadly positive improvements in SAS. Disease traits such as allergic disease and Dyslip showed relative R^2^ gains of 6.8% and 12.8% with SBayesCO-eQTL, and 4.1% and 10.2% with SBayesCO-pQTL, respectively.

Blood cell count traits also showed consistent improvements, including PLC (1.8%–3.5%), neutrophil count (NeuC; 5.0%–5.8%), and red blood cell (erythrocyte) count (RBC; 3.2%–3.7%). VitD remained an exception, with prediction accuracy decreasing under SBayesCO-eQTL (−9.6%) but increasing under SBayesCO-pQTL (5.7%).

These results indicate that incorporating molQTL effect sizes into polygenic prediction provides the greatest benefit for trans-ancestry prediction in the South Asian population. This likely reflects the relatively closer genetic relationship between South Asian and European populations, which may allow EUR-trained molQTL effects to transfer more effectively. In contrast, prediction in more genetically divergent populations, such as AFR and EAS, showed limited or inconsistent gains, suggesting reduced transferability of EUR-trained effects at both molecular phenotype and complex trait levels across ancestries.

### SBayesCO prioritises high-confidence SNPs in regulatory genomic regions

To better understand how different modelling strategies leverage molQTL information, we analyzed the functional enrichment of posterior inclusion probability (PIP) SNPs (moderate: PIP > 0.1; high: PIP > 0.5) across all 11 traits, stratified into four categories: genic-eQTL, genic-pQTL, genic-epQTL (SNPs annotated as both eQTL and pQTL), and intergenic regions (Fig. 5; Supplementary Table 6). These categories are defined at the SNP level based on molQTL annotations rather than representing subsets of genes. Functional enrichment was defined as the ratio between the proportion of PIP-selected SNPs and the genome-wide proportion of SNPs within each category, thereby accounting for differences in annotation size.

**Fig. 5.**
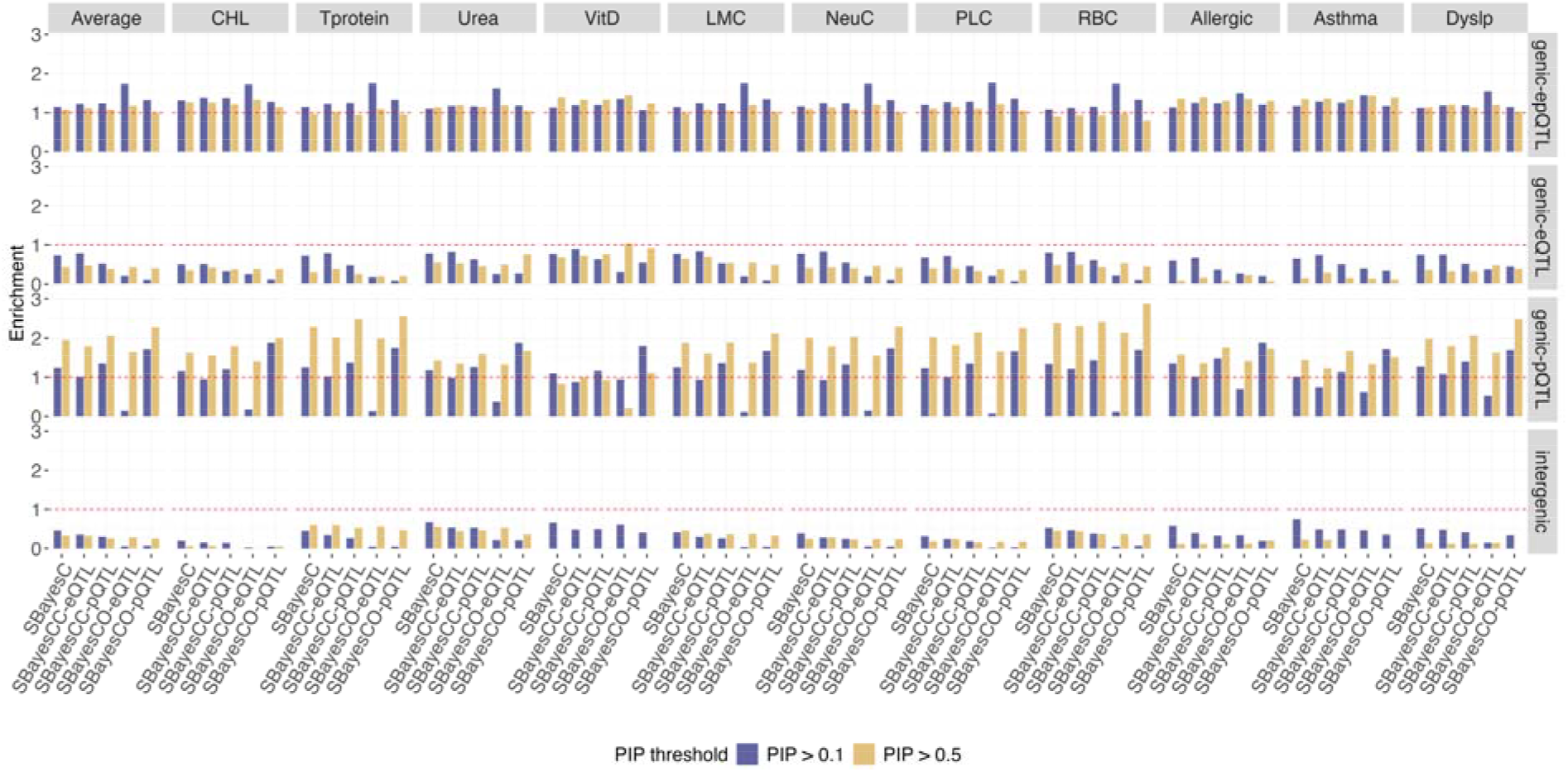
Functional enrichment of regulatory annotations across methods for blood-related traits based on moderate and high PIP SNPs. Enrichment of SNPs with moderate (PIP > 0.1 and high (PIP > 0.5) PIP across functional annotation categories. Enrichment is defined as the ratio between the proportion of PIP-selected SNPs and the proportion of all SNPs within each functional category. Four functional categories are considered: genic-eQTL, genic-pQTL, genic-epQTL (overlapping eQTL and pQTL annotations), and intergenic regions. Results are shown for five models across 11 UK Biobank traits, with an additional “Average” panel summarising enrichment patterns aggregated across all traits. Traits are grouped into blood biomarkers (CHL, Tprotein, Urea, VitD), blood cell counts (LMC, NeuC, PLC, RBC), and immune-related diseases (Allergic, Asthma, Dyslip) (Supplementary Table 1).

On average across the 11 traits, incorporating molQTL information progressively concentrated PIP-selected SNPs into regulatory genic regions while depleting intergenic signals. At PIP > 0.1, the intergenic proportion decreased from 5.6% under the annotation-free baseline to 3.7%–4.3% under SBayesCC, and further to <1% under SBayesCO, corresponding to a drop in the enrichment from 0.5 (SBayesC) to 0.3–0.4 (SBayesCC) and to 0.05–0.06 under SBayesCO. Similar patterns were observed at PIP > 0.5, where intergenic SNPs remain depleted (proportion 3% and enrichment 0.3 across models), while regulatory regions dominated high-confidence signals.

Beyond intergenic regions, modelling strategies also differed in how regulatory signals were redistributed across functional categories, and these differences were consistently reflected at both modest and high PIP thresholds. At PIP > 0.1, on average, SBayesC enriched PIP-selected SNPs in genic regions, with enrichment of 0.7 in genic-eQTL, 1.1 in genic-epQTL, and 1.2 in genic-pQTL regions. Incorporating binary molQTL annotations in SBayesCC strengthened this pattern, with SBayesCC-eQTL increasing enrichment in genic-eQTL and genic-epQTL regions (enrichment 0.8 and 1.2), and SBayesCC-pQTL preferentially enriching genic-pQTL regions (enrichment 1.3). Effect-size-based modelling in SBayesCO further amplified regulatory prioritisation, with genic-epQTL enrichment reaching 1.7 under SBayesCO-eQTL and genic-pQTL enrichment reaching 1.7 under SBayesCO-pQTL. Similar patterns were observed at PIP > 0.5, indicating that SBayesCO preserves regulatory prioritisation among the highest-confidence SNPs.

Several traits showed method-specific changes in enrichment patterns as the PIP threshold increased from 0.1 to 0.5, reflecting a transition from modest-to high-confidence signals. For blood cell traits such as RBC and PLC, enrichment in genic pQTL regions increased consistently across models but was most pronounced under SBayesCO-pQTL: genic-pQTL enrichment rose from approximately ∼1.6–1.7 at PIP > 0.1 to ∼2.3–2.9 at PIP > 0.5, compared with ∼2.3–2.4 under SBayesC and SBayesCC-pQTL. This pattern indicates that effect-size-based modelling further concentrates the highest-confidence signals into protein-regulatory loci beyond annotation-based approaches alone.

These results indicate that although eQTLs and pQTLs capture different regulatory signals, effect-size-based modelling promotes convergence of SNP prioritisation towards shared regulatory loci. Notably, even when trained on a single molQTL type, SBayesCO implicitly favoured variants located in overlapping regulatory regions, suggesting its potential for joint molQTL integration. This refined prioritisation is consistent with the observed improvements in prediction accuracy, suggesting that concentrating signals within regulatory regions improves the power of causal variant identification and effect estimation.

### Locus-specific examples of SNP prioritization using SBayesCO

Although SBayesCO was developed primarily to improve polygenic prediction, its bivariate Bayesian framework of joint modelling of GWAS and molQTL effects is closely related to SNP prioritisation. Across multiple loci, integrating molQTL-derived effect sizes yields more concentrated posterior inclusion probability (PIP) profiles and reduces ambiguity in regions where GWAS analysis or baseline Bayesian models were unable to distinguish correlated variants (Fig. 6).

**Fig. 6.**
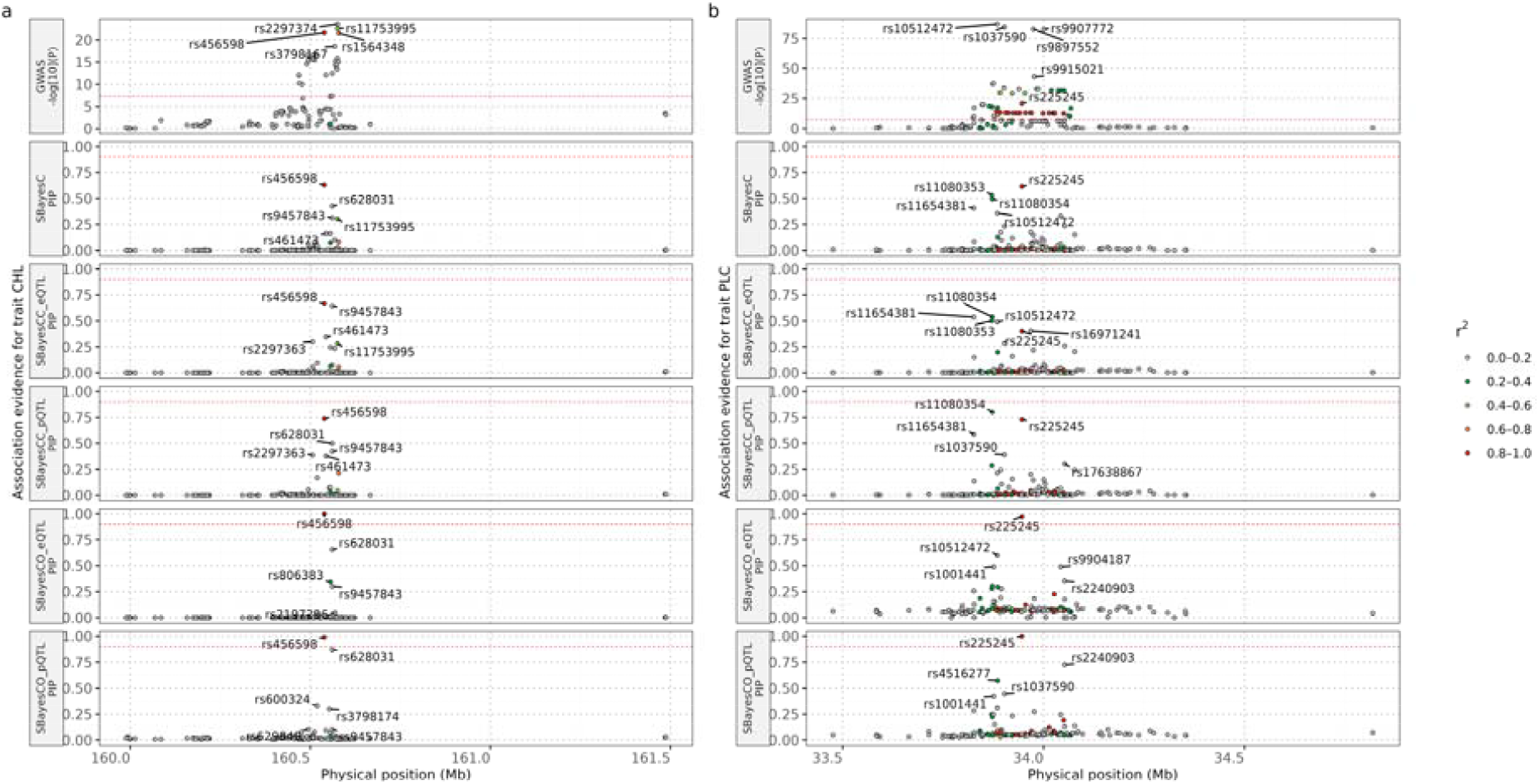
Comparison of fine-mapping results across methods for total cholesterol (CHL) and platelet count (PLC) in two genomic regions. LocusZoom plots (top row) of the two-sided –log10(P value) from marginal GWAS analyses for CHL (a) and PLC (b) across base pair positions. The horizontal dashed line indicates the genome-wide significance threshold (P=5×10□□). Point colours denote LD (R^2^) with the fine-mapped variant in each region. Rows below each GWAS panel show posterior inclusion probabilities (PIPs) across the same genomic regions, obtained from five Bayesian fine-mapping approaches: SBayesC, SBayesCC-eQTL, SBayesCC-pQTL, SBayesCO-eQTL, and SBayesCO-pQTL. SNPs with PIP > 0.9 are highlighted as putative causal variants. Compared with SBayesC and SBayesCC, SBayesCO yielded higher PIPs for complex traits at SNPs with regulatory roles (high PIPs for eQTL or pQTL effects), indicating improved resolution of candidate causal variants for both traits.

For CHL, the marginal GWAS signal formed a broad association peak centred around rs456598 (P=2×10^-22^), with extensive LD obscuring the identity of the causal variant. SBayesC produced moderate support to rs456598 (PIP=0.63), and SBayesCC modestly increased this support (PIP=0.67 with eQTL and 0.74 with pQTL), but substantial posterior mass remained distributed across neighbouring SNPs. In contrast, both SBayesCO-eQTL (PIP = 1.00) and SBayesCO-pQTL (PIP = 0.99) sharply prioritise rs456598 while down-weighting LD-correlated variants that appear equally plausible under traditional frameworks. The SNP rs456598 is located proximal to the promoter of SLC22A1 (3.9 kb upstream; HaploReg v4.2^41^), a gene with strong prior evidence linking it to serum lipid traits, including total cholesterol. Multiple GWAS have associated *SLC22A1* variants with total cholesterol, LDL□cholesterol, and triglycerides, and functional studies indicate a role for *SLC22A1* in hepatic lipid metabolism^42,43^.

A similar pattern was observed for PLC, where several variants, including rs225245, rs11080354 and rs10512472, exhibited comparable marginal association strengths within a highly correlated LD cluster. Notably, rs225245 lies in an intronic region of *AP2B1*, a gene previously implicated in platelet count through GWAS and integrative analyses such as TWAS^44^ and SMR^43^. Neither SBayesC nor SBayesCC was able to reliably distinguish among these candidates, whereas SBayesCO-eQTL (PIP = 0.97) and SBayesCO-pQTL (PIP = 1.00) consistently prioritized rs225245 as the most likely causal variant. This refinement of PIP profiles suggests that molQTL effect size information helps resolve regulatory heterogeneity that is not be identifiable using GWAS data alone.

Collectively, these examples show that although SBayesCO was motivated by improvements in polygenic prediction, the bivariate framework incorporating molQTL effect sizes facilitate SNP prioritization. By directly integrating functional effect-size information, SBayesCO reduces posterior uncertainty and prioritizes biologically plausible candidate variants, thereby improving interpretability across traits and genomic regions.

## Discussion

In this study, we introduced SBayesCO, a Bayesian framework that integrates GWAS summary statistics and molQTL information through modelling SNP joint effect sizes on both complex trait and molecular phenotypes to improve polygenic prediction. Unlike baseline models such as SBayesC, which ignores functional information, or SBayesCC, which relies on binary annotations, SBayesCO incorporates molQTL effect sizes as regulatory evidence, allowing SNP effect distributions to be informed by the genetic covariances between the complex trait and thousands of genes or proteins. Across simulations and applications to 11 complex traits using both eQTL and pQTL data, SBayesCO consistently improved prediction accuracy compared to the baseline models (Fig. 2-3).

The improved performance of SBayesCO likely arises from its ability to exploit the relationship between molQTL effects and complex trait effects. In genomic regions where SNP effects on the molecular phenotype and the complex trait are directionally aligned, large molecular effects provide informative signals for prioritizing SNPs that also have large effects on the trait. As a result, SNPs with strong molecular effects in such regions are more likely to receive large estimated trait effects, thereby improving polygenic prediction accuracy. However, the correlation between molecular and complex trait effects is estimated from a relatively small subset of SNPs affecting both traits, which may result in estimation noise and occasionally lead to slightly lower predictive performance than SBayesC. This limitation could potentially be mitigated by clustering genes with similar genetic covariance with the trait and estimating shared genetic covariance across all cis-SNPs within each gene cluster.

Compared to existing methods, SBayesCO offers a more biologically informed and flexible framework for integrating molQTL data. Traditional approaches,, such as SBayesC, ignore functional annotations, thereby distributing genetic signals more diffusely across the genome. Existing methods that do incorporate functional data, such as LDpred-funct^21^, AnnoPred^19^, PolyFun^20^, and SBayesRC^23^, typically treat functional relevance as categorical annotations that influence SNP effect variances or inclusion probabilities. However, when molQTL are encoded as binary annotations, all SNPs associated with gene expression are treated as equally relevant, regardless of whether the corresponding gene influences the trait. This will introduce noise and dilute signals arising from true expression mediation. In contrast, SBayesCO directly models heterogeneity in effect-size distributions, using both the direction and magnitude of molQTL effects to differentially weight SNPs based on regulatory relevance.

The significant advantage of SBayesCO over SBayesCC provides general guidance on how quantitative genomic annotations, beyond molQTL, can be effectively integrated into polygenic prediction models. For example, emerging DNA foundation models^45-47^ now generate extensive AI-predicted regulatory variant effect scores, often with tissue or cell-type resolution. Recent studies have begun using these predictions as genomic annotations to improve polygenic prediction and fine-mapping. Our findings suggest that integrating such quantitative regulatory effect predictions through multivariate effect-size modelling, rather than treating them as binary or categorical annotations, may be a more powerful strategy to fully realise their potential.

Comparisons between eQTL- and pQTL-integrated models provide additional mechanistic insights. Across traits, SBayesCO-pQTL outperformed SBayesCO-eQTL in prediction accuracy (Fig. 3), and high-PIP SNPs were more enriched in post-transcriptional regulatory regions (Fig. 5). Consistent with protein abundance acting closer to complex traits than gene expression, these results suggest that pQTLs capture non-redundant biological signals beyond transcriptional control^48^. Interestingly, even when models were trained on a single molQTL type, SBayesCO consistently prioritized SNPs supported by multiple regulatory signals (Fig. 5), indicating that shared regulatory signals can be captured implicitly through effect-size modelling. These results motivate future efforts to unify multiple molQTL sources, such as eQTL, pQTL, and methylation QTL, within a single multivariate modelling framework^49^.

While SBayesCO shows robust performance across traits and annotations, several limitations remain. First, it assumes no sample overlap between GWAS and molQTL datasets. Although this assumption is reasonable for eQTL resources, it is known to be violated for the pQTL data analysed here, where partial sample overlap with the GWAS cohorts exists^37^. Such overlap could, in principle, induce correlations between GWAS and pQTL effect estimates and inflate posterior inclusion probabilities. A more explicit modelling of sample overlap represents an important direction for future methodological development^50^. Second, the present study adopts BayesC as the baseline model and restricts analyses to HapMap3 SNPs in order to provide a controlled and computationally tractable setting for evaluating the incremental contribution of molQTL-integrated effect size priors. While this design facilitates interpretation, the limited flexibility of the baseline model and the reduced SNP density may constrain the extent to which molQTL information can be exploited. Although our results demonstrate that integrating molQTL can aid SNP prioritization, full fine-mapping applications would require modelling all imputed or sequence variants. Future work could extend this framework to more flexible baseline models, such as SBayesR^51^, and to sequence-level data, potentially incorporating multiple layers of functional annotation as demonstrated in recent large-scale studies (e.g. OPERA^49^). Third, inference via Gibbs sampling, although stable, is computationally intensive; faster alternatives, such as variational Bayes^52^ or scalable MCMC methods^53^, could enhance efficiency. Fourth, SBayesCO currently treats eQTL and pQTL separately and does not leverage tissue-specific, ancestry-aware^54,55^, or trans-QTL effects^56^, which are known to explain a substantial fraction of gene expression variance but whose summary statistics are typically unavailable in large-scale studies. Integrating resources like GTEx^57^, with more complex hierarchical^58^ and mixture priors^8,59^ would improve generalisability.

In summary, SBayesCO introduces an effect-size–based strategy for integrating molQTL data into polygenic models, leveraging both the direction and magnitude of molQTL effects. This approach improves polygenic prediction while enhancing functional interpretability. This framework provides a general and extensible approach for incorporating functional genomic data into polygenic prediction and SNP prioritization across diverse molecular datasets and traits.

## Methods

### Ethics approval

The University of Queensland Human Research Ethics Committee B (2011001173) provides approval for analysis of human genetic data used in this study on the high-performance cluster of the University of Queensland.

### SBayesCO Model

We start by describing the individual-level model and then introduce its variant based on the summary statistics only. Relevant technical details are provided in Supplementary Notes 1. Let **y** be the *n*_*t*_ × 1 phenotype vector for the complex trait, **W**_*k*_ be the *n*_*m*_ × 1 expression vector for the *k*-th molecular phenotype, **G** be the *n*_*t*_ ×*P*_*int*_ genotype matrix for SNPs in the intergenic region, **X**_*k*_ and **Z**_*k*_ be the *n*_*t*_ × *p*_*k*_ and *n*_*m*_ × *p*_*k*_ genotype matrices for SNPs in the *k*-th genic region in the complex trait sample and molecular phenotype sample, respectively, with *p*_*int*_ being the number of intergenic SNPs, and *p*_*k*_ being the number of cis-SNPs for the *k*-th molecular phenotype. As we consider only the cis region for each molecular phenotype (i.e., 2 Mb around the mid-position of the gene), *p*_*k*_ ≪ *p*_*int*_ < *p* the total number of SNPs. In addition, since the molecular phenotypes are often measured in a different sample from that for the complex trait, *n*_*t*_ and *n*_*m*_ can be different, so do **X**_*k*_ and **Z**_*k*_. We assume that **y**,**w**_*k*_, and each column of **G, X**_*k*_ and **Z**_*k*_ have been standardised with zero mean and unit variance. Our bivariate model that combines the complex trait and the *k*-th molecular phenotypes, accounting for both intergenic and genic SNP effects, is

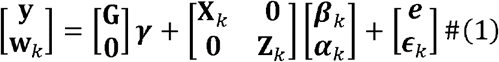

where ***γ*** is the vector of trait effects of intergenic SNPs, **β**_*k*_ is the vector of trait effects of genic SNPs pertaining to the *k*-th molecular phenotype, ***α***_*k*_ is the vector of these SNP effects on the molecular phenotype, **e** and ***ϵ***_*k*_ are corresponding residuals for the complex trait and the molecular phenotype. The residual vector 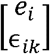 for the *i*-th sample follows a bivariate normal distribution 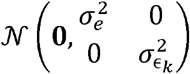. In this model, we can treat the SNP *j* in the intergenic region as a special bivariate case where ***α***_*k*_ = **0**.

Extending this formulation to *K* molecular expression gives a multivariate regression model that jointly incorporates the complex trait and all molecular phenotypes:

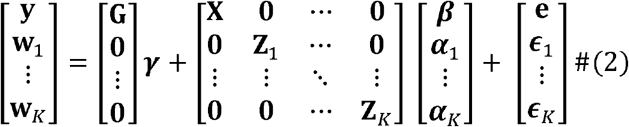

Under the assumption of independence between molecular phenotypes and considering only cis-region of each molecular phenotype, this multivariate model collapses to a set of independent bivariate models as shown in Eq (1).

#### Priors for intergenic SNP effects

The effect of SNP*j* in the intergenic region follows a mixture of a point mass at zero with probability *π*_*γ*_ and a univariate normal distribution with probability 1 − *π*_*γ*_ :

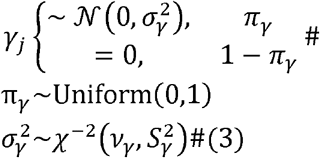

This is identical to the priors in BayesC^39^.

#### Priors for genic SNP effects

For SNPs in the genic region of the *k*-th molecular phenotype, each have two effects: the effect on the trait *β*_*jk*_ and the effect on the molecular phenotype *α*_*jk*_. The prior *β*_*jk*_ for *α*_*jk*_ or is assumed to be a mixture with a point mass at zero and a univariate normal distribution:

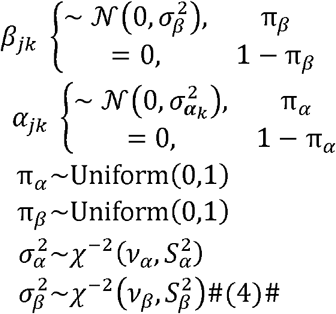

This prior is referred to as the “either in either out” (EIEO) prior, as it allows the SNP to have a non-zero effect on either complex trait or molecular phenotype, or both, thereby accounting for both trait-specific and pleiotropic effects.

The covariance between molQTL effects at the same SNP for any two molecular phenotypes is

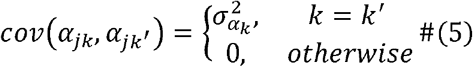

where independence of molQTL effects is assumed between two distinct molecular phenotypes. The covariance between effects at the same SNP *j*. for molecular phenotype and complex trait is

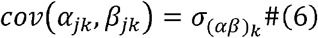

For ease of computation, we use an alternative parameterisation for the above prior. Let 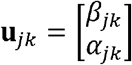 and 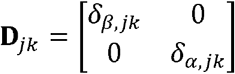 where δ_*β,jk*_ *∼Bernoulli(π*_*β*_) and δ_*α,jk*_ *∼Bernoulli(π*_*α*_) are indicator variables for the complex trait effect and molecular phenotype effects, respectively. Then we can write the EIEO prior as

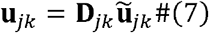

where 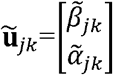 is a latent SNP effect pair following a bivariate normal distribution:

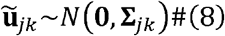

where the variance-covariance matrix is assumed to follow an inverse-Wishart distribution:

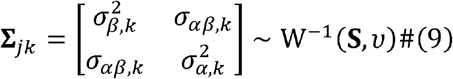

With the scale matrix 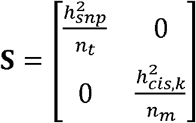 and the degrees of freedom *υ* = 4.

Therefore, the genetic covariance between the complex trait and molecular phenotype is modelled through the covariance between the latent effects at the SNPs in the cis region.

If there are K^′^ overlapping genes at the SNP for gene *k*^′^, ⃛, *K*^′^, the SNP will have *K*^′^ molQTL effects (*α*_*jk*_′, ⃛,α_*jK*′_ ). To maintain the bivariate structure in the model, we partition the complex trait β_*jk*_ effect into additive components corresponding to the molQTL effects

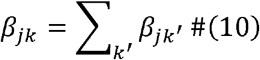

Therefore,

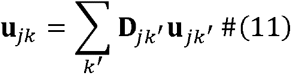

where each **u**_*jk*_ ^′^ has an independent prior, as in Eq (9).

#### Heritability estimation

SNP-based heritability is defined as the variance in the complex trait explained by all SNPs (assuming unit trait variance):

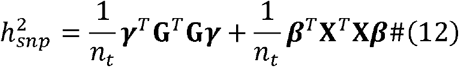

Cis-heritability for the *k*-th molecular phenotype is defined as the variance in the *k*-th molecular phenotype explained by the predicted genetic component of the *k*-th molecular phenotype (assuming unit variance for molecular phenotype):

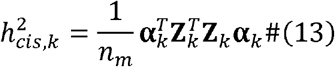

#### Extensions to summary statistics

According to ref^23^, we can transform our individual-level model into a summary-level model:

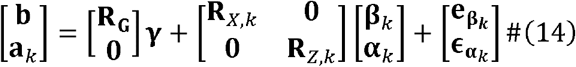

where **b** is the vector of GWAS marginal effect size, **a**_*k*_ is the vector of molQTL marginal effect size in the *k* -th molecular phenotype, **R**_**G**_, **R**_**X**_, and **R**_**Z**_ are the corresponding LD correlation matrices, residual vector 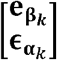 follows a multiple normal distribution 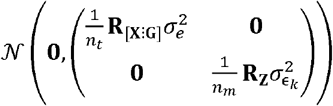, [**X**: **G**] is the combined genotype matrix where SNPs from both genic and intergenic region. Notably, in our summary-level model, for complex traits, we partitioned the whole genome into 591 LD blocks with a minimum width of 4 cM^23^. In contrast, for molecular traits, a molecular-specific LD matrix is generated. As a result, **R**_**X**_ and **R**_**Z**_ will differ, reflecting the distinct LD structures used for complex and molecular traits.

Here, eigen-decomposition is applied to genome-wide and cis-region LD matrices to avoid potential convergence problems caused by sparse LD matrices^23^. For each LD block, 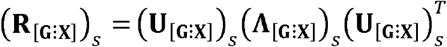, and for the *k*-th molecular phenotype, 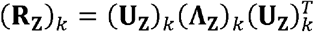, where 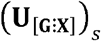 and 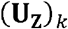 are the corresponding block-wise and molecular-wise matrices of eigenvalues(**⋀**_[**G: X**]_ )_s_ and (**⋀Z**)_*k*_ are the corresponding block-wise and molecular-wise diagonal matrices of eigenvalues. By multiplying both sides of the equation (3) by 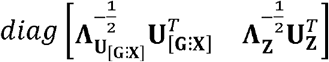 a low-rank summary model can be further constructed as

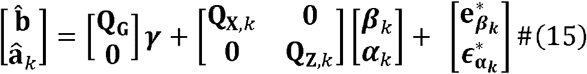

Where 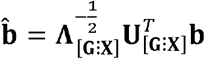 and 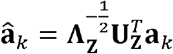 are linear combinations of marginal GWAS SNP effects and marginal molecular SNP effects, separately. After applying eigen-decomposition methods, the dimension of 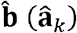 will be less than or equal to the dimension of 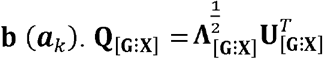and 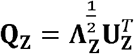 are the corresponding new coefficient matrices and the new residuals 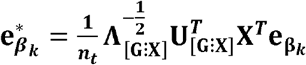 and 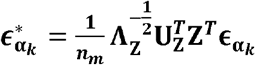, residual vector 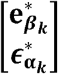 follows a multiple normal distribution 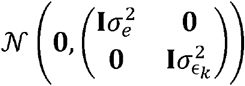.

Correspondingly, SNP-based heritability is

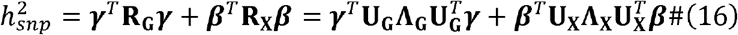

and cis-heritability for the *k*-th molecular phenotype is

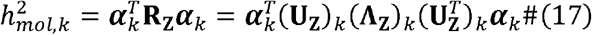

A total of 3,000 MCMC samples were drawn, with the initial 2,000 iterations treated as burn-in and the final 1,000 retained for posterior estimation.

### SBayesC and SBayesCC

SBayesC is a genome-wide Bayesian framework that utilises GWAS summary statistics to infer joint SNP effects. In this framework, SNP effect sizes are characterised using a two-part mixture: either taking a null effect with probability 1-*π*, or drawn from a Gaussian distribution with probability *π*^39^. To avoid convergence issues, eigen-decomposition was applied to genome-wide LD matrices.

SBayesCC is a genome-wide Bayesian framework designed for heritability estimation and polygenic prediction by jointly evaluating GWAS summary data and functional annotation profiles. This method builds upon the SBayesRC^23^ framework, adopting a two-part prior distribution that models SNP effects as either null or drawn from a normal distribution regulated by a sparsity-controlling parameter *π*. Different molQTL annotations are incorporated to assign elevated prior probabilities to SNPs located in biologically meaningful genomic regions.

### Prediction accuracy estimation

To evaluate predictive performance, a five-fold cross-validation framework was applied to the UK Biobank dataset, involving 341,809 unrelated individuals of European ancestry. For each fold, 80% of the samples were used to train the model, while the remaining 20% served as the validation set. GWAS summary statistics were computed using PLINK2 software^60^, adjusting for covariates, including sex, age, and the top 10 genetic principal components. Trait-specific regression models were employed, with linear regression applied to continuous outcomes and logistic regression used for binary traits.

### Simulation

To assess the performance of the SBayesCO method, we simulated gene expression levels and a quantitative trait phenotype using genotypes from the UK Biobank, limited to HapMap 3 SNPs on chromosome 22 with 12,159 SNPs in total and five random subsamples (5K, 10K, 50K, 100K, and 300K) from 348,501 unrelated European individuals. Gene information was obtained from hg19 GENCODE (v40) within a 50-kilobase (KB) window around the midpoint of each gene. Using the genotypes and gene information, two simulation scenarios—causal and realistic settings—were applied to generate gene expression levels and complex trait phenotypes, with 20 independent replicates for each scenario (Fig. 1a). The total cis heritability 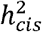 was set to be 0.5 for all simulation scenarios.

In the causal scenario, where genetic variants affect the complex trait via gene regulation, we randomly selected *M*_*cau*_ = 50 causal genes and, on average, 10 causal variants for each gene. The exact number of causal variants *C*_*cau,k*_ for each gene *k* followed a Poisson distribution *C*_*cau,k*_ ∼ *Poission*(*λ* = 10) and the effect size α_*jk*_ for each causal variant *j* was drawn from a normal distribution 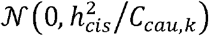, where 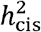 denotes the cis-heritability of gene expression attributable to variants within the cis region of a gene. The *k*-th gene expression levels **w**_*k*_ were simulated based on an additive model **w**_*k*_ = **Z**_*k*_ ***α***_*k*_ + **ϵ**_*k*_ with environmental noise **ϵ**_*k*_ drawn from a multivariate normal distribution 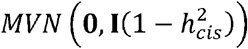. The *k*-th gene effect θ_*k*_ was simulated from a normal distribution 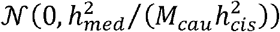,, where 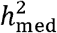 represents the mediated heritability, defined as the proportion of phenotypic variance explained by genetic effects mediated through gene expression. Together with expression levels simulated in the previous step, the complex phenotypes **y** were generated based on an additive mode **y** = **∑**_*k*_ **Z**_*k*_***α***_*k*_*θ*_*k*_+**e** with environmental noise **e** drawn from a multivariate normal distribution 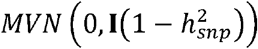. Here, 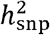 denotes the total SNP heritability of the trait, defined as the proportion of phenotypic variance explained by all genetic variants. Both mediated heritability 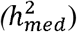 ) and total SNP heritability 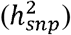 were set to 0.1.

In the realistic scenario, we examine SNP sets from both genic and intergenic regions, focusing on three groups of genes: the causal gene set, the pleiotropic gene set (where cis-SNPs influence both the complex trait and gene expression separately), and the extra gene set (where cis-SNPs solely regulate gene expression). As described above, 50 causal genes were simulated in the genic region. In the pleiotropic scenario, similar to those in the causal model, we simulated *M*_*ple*_ = 50 gene expression levels and the exact number of causal variants *C*_*cau,k*_ for each gene *k* followed a Poisson distribution *C*_*ple,k*_ ∼ *Poission*(*λ* = 10). Additionally, *M*_*non*_ = 50 extra genes with a total of *M*_*non*_ ∑_*k*_ *C*_*non,k*_ causal variants for gene expression only were generated, with each variant effect *α*_*kj*_ drawn from a normal distribution 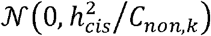, where the exact number of causal variants for each gene *C*_*non,k*_ ∼ *Poission*(*λ* = 10). In the intergenic region, a total of *C*_*int*_ = 1000 causal variants were selected. Together with all pleiotropic variants from the genic region, the effect size of each causal varian *l, β*_*l*_ is generated from a normal distribution 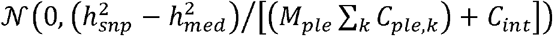. Finally, the complex phenotypes **y** were 96 simulated from the equat **y** = **Xβ + ∑**_*k*_ **Z**_*k*_***α***_*k*_*θ*_*k*_+**e** with environmental noise **e** drawn from a multivariate normal distribution 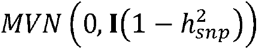. Accordingly, the *k*-th gene effect *θ*_*k*_ from pleiotropic and extra gene sets was 0. Mediated heritability 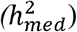 and total SNP heritability 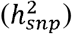 599 were set to 0.05 and 0.1, respectively.

For each of the 20 replicates in the scenarios above, centred and scaled genotypes were used. Gene expression levels and trait phenotypes were centred. Simple linear regression was employed to calculate GWAS and eQTL marginal summary statistics. Two genotype datasets, consisting of 1K and 50K individuals, were randomly selected from unrelated European samples. Then, they were used to generate external gene expression levels with the same eQTL settings mentioned above for each model. We further generated independent validation and LD samples from the remaining unrelated European individuals, with a sample size of 10K for validation and 5K for LD. For each simulation scenario, SBayesC (as benchmark) and SBayesCO were used. To assess prediction accuracy, we generated a polygenic risk score for each sample using the genotypes from the validation datasets and the genetic effects estimated from each method. The prediction was calculated by regressing the true simulated phenotype of the complex trait against that predicted from each method.

#### Independent traits and genotypes

We selected 11 independent traits from the UK Biobank dataset, restricted to 341,809 unrelated individuals of European descent (Supplementary Table 1). For continuous phenotypes, extreme values beyond ±7 standard deviations from the mean were excluded, followed by a rank-based inverse normal transformation, applied separately within European ancestry and sex groups. Genotype quality control involved removing multi-allelic variants, as well as SNPs with imputation information scores < 0.3, hard-call genotype probability < 0.9, minor allele frequency (MAF) < 0.01, Hardy–Weinberg equilibrium p-values < 10□□, or missingness rates > 1%. After filtering and restricting to 1 million HapMap 3 SNPs, approximately 1,154,522 SNPs high-quality SNPs were retained for downstream analysis. To define the cis-region for a gene, we mapped SNPs to a flanking window of 1MB on both sides of the gene midpoint position based on GENCODE (v40, https://www.gencodegenes.org/human/release_40.html) under the hg19 genome build (50KB was used as the flanking window size in simulations).

#### Expression data

After retaining SNPs common to both the 1M HapMap3 SNP set and the LD reference panel, a total of 8,705 genes with 834,893 cis-eQTLs and 2856 proteins with 797,070 cis-pQTLs remained (Supplementary Table 2).

## Data Availability

Individual-level genotype and phenotype data were obtained from the UK Biobank resource under application number 12505.
These data are available through application to the UK Biobank (https://www.ukbiobank.ac.uk/).
Summary statistics used in this study were obtained from previously published studies, including the eQTLGen consortium and the UK Biobank Pharma Proteomics Project (UKB-PPP) study, and are available from the original publications or their respective repositories.

## Data availability

No data were generated in the present study. The eQTL summary statistics are publicly available at https://www.eqtlgen.org/. The pQTL summary statistics are available at https://www.synapse.org/Synapse:syn51364943/. The UK Biobank data are from the UK Biobank resource at https://www.ukbiobank.ac.uk/enable-your-research/about-our-data/.

## Code availability

For plink2.0, see https://www.cog-genomics.org/plink/2.0/. For GCTA-COJO, see https://yanglab.westlake.edu.cn/software/gcta/#COJO. For SBayesRC, see https://cnsgenomics.com/software/gctb/#SBayesRCTutorial. SBayesCO is implemented in an R package available at https://github.com/ShouyeLiu/SBayesOmics and in the publicly available software BayesOmics at https://github.com/ShouyeLiu/BayesOmics.

## Acknowledgements

We thank Suet Ching Phoebe Hui for helpful discussions and insightful comments. We also thank the Research Computing Centre (RCC) Infrastructure Team in the University of Queensland for their support in this research. This research was supported by the Australian National Health and Medical Research Council (1177268 and 2043111) and the Australian Research Council (FL180100072). P.M.V. acknowledges funding from the European Research Council (grant 101198904). This study makes use of data from the UKB (project ID 12505).

## Author contributions

J.Z. conceived and designed the study. J.Z. and S.L. developed the methods and algorithms. S.L. conducted all analyses with the assistance or guidance from M.G., H.C., J.Y., P.M.V., and J.Z. S.L. developed the C++ software and R package with the guidance from J.Z. S.L. and J.Z. wrote the manuscript with the participation of all authors. All the authors approved the final version of the manuscript.

## Competing interests

The authors declare no competing interests.

## Additional information

Supplementary information is available for this paper at the link.

## Notes

### Competing Interest Statement

The authors have declared no competing interest.

### Author Declarations

The North West Multi-centre Research Ethics Committee gave ethical approval for the UK Biobank study. This research used individual-level genotype and phenotype data from the UK Biobank resource under application number 12505. Additional summary statistics were obtained from previously published studies, including the eQTLGen consortium and the UKB-PPP study.

