## Supplementary Notes for "Joint Bayesian modelling of molecular QTL and GWAS effects improves polygenic prediction for complex traits"

**1. Supplementary Note 1 – MCMC sampling strategy in the individual-level model and summary-level model**

**MCMC sampling in the individual-level model.** MCMC sampling is used to draw posterior inferences from the model parameters. The joint distribution of data and all parameters in the individual-level model is

$$\begin{aligned} f\left( \mathbf{y},\mathbf{w},\boldsymbol{\beta},\boldsymbol{\alpha},\boldsymbol{\Sigma},\boldsymbol{\pi},\sigma_{e}^{2},\boldsymbol{\sigma}_{\boldsymbol{\epsilon}}^{2} \right)&\propto\prod_{k=1}^{K} exp\left\{ -\frac{1}{2}\sum_{i=1}^{n_{t}} \left[ \mathbf{A}^{T}\mathbf{E}^{-1}\mathbf{A} \right] \right\}\# \\ &\times\prod_{k=1}^{K} \prod_{j=1}^{p_{k}} \left| \boldsymbol{\Sigma}_{jk} \right|^{-\frac{1}{2}}exp\left\{ -\frac{1}{2}\mathbf{u}_{jk}^{T}{\boldsymbol{\Sigma}_{jk}}^{-1}\mathbf{u}_{jk} \right\} \\ &\times\prod_{k=1}^{K} \frac{\left| \mathbf{S}_{\Sigma} \right|^{v_{\boldsymbol{\Sigma}}/2}\left| \boldsymbol{\Sigma}_{jk} \right|^{-\left( v_{\Sigma}+2+1 \right)/2}exp\left\{ -tr\left( \mathbf{S}_{\Sigma}\boldsymbol{\Sigma}_{k}^{-1} \right) \right\}}{2^{{2v_{\Sigma}}/2}\Gamma\left( {v_{\Sigma}}/2 \right)} \\ &\times\prod_{k=1}^{K} \frac{exp\left( {-\nu_{\epsilon,k}S_{\epsilon,k}^{2}}/{2\sigma_{\epsilon_{k}}^{2}} \right)}{\left( \sigma_{\epsilon_{k}}^{2} \right)^{1+{\nu_{\epsilon,k}}/2}} \\ &\times\prod_{j=1}^{p_{int}} \left( 2{\pi\sigma}_{\gamma}^{2} \right)^{-\frac{1}{2}}exp\left[ \frac{\gamma_{j}^{2}}{2\sigma_{\gamma}^{2}} \right] \\ &\times\frac{\exp\left( {-\nu_{\gamma}S_{\gamma}^{2}}/{2\sigma_{\gamma}^{2}} \right)}{\left( \sigma_{\gamma}^{2} \right)^{1+{\nu_{\gamma}}/2}} \\ &\times\frac{\exp\left( {-\nu_{e}S_{e}^{2}}/{2\sigma_{e}^{2}} \right)}{\left( \sigma_{e}^{2} \right)^{1+{\nu_{e}}/2}}\#(S1) \end{aligned}$$

Where $\mathbf{E}=\left[ \begin{matrix} \sigma_{e}^{2} & 0 \\ 0 & \sigma_{\epsilon_{k}}^{2} \end{matrix} \right]$, $\mathbf{A}=\left[ \begin{matrix} y_{i} \\ w_{ik} \end{matrix} \right]-\sum_{j=1}^{p_{int}} \left[ \begin{matrix} G_{ij} & 0 \\ 0 & 0 \end{matrix} \right]\left[ \begin{matrix} \gamma_{j} \\ 0 \end{matrix} \right]\boldsymbol{-}\sum_{j=1}^{p_{k}} \left[ \begin{matrix} \left( \mathbf{X}_{k} \right)_{ij} & 0 \\ 0 & \left( \mathbf{Z}_{k} \right)_{ij} \end{matrix} \right]\mathbf{D}_{jk}\mathbf{u}_{jk}$.

Suppose $\eta_{j}$ is the indicator variable to the distribution membership of $\gamma_{j}$. The full conditional distribution of $\gamma_{j}$ is

$$\begin{aligned} f\left( \gamma_{j}|\boldsymbol{y},\boldsymbol{\gamma}_{-j},\eta_{j}=1,\sigma_{\gamma}^{2},\sigma_{e}^{2} \right)&\propto exp\left\{ -\frac{1}{2\sigma_{e}^{2}}\left( \tilde{\mathbf{y}}-\mathbf{G}_{j}\gamma_{j} \right)^{T}\left( \tilde{\mathbf{y}}-\mathbf{G}_{j}\gamma_{j} \right) \right\}exp\left\{ -\frac{\gamma_{j}^{2}}{2\sigma_{\gamma}^{2}} \right\}\# \\ &\propto N\left( \frac{r_{j}}{C_{j}},\frac{\sigma_{e}^{2}}{C_{j}} \right)\#(S2) \end{aligned}$$

Where

$$\begin{aligned} \tilde{\mathbf{y}}\mathbf{&=y}-\sum_{k} \mathbf{X}_{k}\boldsymbol{\beta}_{k}\mathbf{-}\sum_{j^{'}\neq j} \mathbf{G}_{j^{'}}\gamma_{j^{'}}\# \\ r_{j}&=\mathbf{G}_{j}^{T}\tilde{\mathbf{y}} \\ C_{j}&=\mathbf{G}_{j}^{T}\mathbf{G}_{j}\boldsymbol{+}\frac{\sigma_{e}^{2}}{\sigma_{\gamma}^{2}}\boldsymbol{\#(}S3\boldsymbol{)} \end{aligned}$$

When $\eta_{j}=0$, $\gamma_{j}=0.$

The full conditional distribution $\eta_{j}$ is

$$\begin{aligned} \Pr\left( \eta_{j}=c|\mathbf{y},\mathbf{G},\boldsymbol{\gamma},\sigma_{\gamma}^{2},\mathbf{X}\boldsymbol{,}\boldsymbol{\beta}\mathbf{,}\sigma_{\beta}^{2},\sigma_{e}^{2} \right)=\frac{f\left( \mathbf{y}\boldsymbol{|}\eta_{j}=c,\mathbf{G},\boldsymbol{\gamma},\sigma_{\gamma}^{2},\mathbf{X}\boldsymbol{,}\boldsymbol{\beta}\mathbf{,}\sigma_{\beta}^{2},\sigma_{e}^{2} \right)f\left( \eta_{j}=c \right)}{\sum_{c^{'}=0}^{1} f\left( \mathbf{y}\boldsymbol{|}\eta_{j}=c^{'},\mathbf{G},\boldsymbol{\gamma},\sigma_{\gamma}^{2},\mathbf{X}\boldsymbol{,}\boldsymbol{\beta}\mathbf{,}\sigma_{\beta}^{2},\sigma_{e}^{2} \right)f\left( \eta_{j}=c^{'} \right)},c=0,1\#\#(S4) \end{aligned}$$

Where $f\left( \mathbf{y}\boldsymbol{|}\eta_{j}=c,\mathbf{G},\boldsymbol{\gamma},\sigma_{\gamma}^{2},\mathbf{X}\boldsymbol{,}\boldsymbol{\beta}\mathbf{,}\sigma_{\beta}^{2},\sigma_{e}^{2} \right)=\int f\left( \mathbf{y}\boldsymbol{|}\eta_{j}=c,\gamma_{j} \right)f\left( \gamma_{j}|\eta_{j}=c,\sigma_{\gamma}^{2} \right)d\gamma_{j}$ above and $f\left( \eta_{j}=1 \right)=\pi_{\gamma}$.

Suppose $\delta_{jk}$ is the indicator variable to the distribution membership of $\mathbf{u}_{jk}$. The full conditional distribution of $\mathbf{u}_{jk}$ is

$$\begin{aligned} f\left( \mathbf{u}_{jk}\mathbf{|y,}\mathbf{w}_{k}\mathbf{,}\mathbf{u}_{-jk}\mathbf{,G,}\mathbf{X}_{k}\mathbf{,}\mathbf{Z}_{k}\boldsymbol{\Sigma,}\delta_{jk}\sigma_{e}^{2}\mathbf{,}\boldsymbol{\sigma}_{\boldsymbol{\epsilon}}^{2} \right)&\propto exp\left\{ -\frac{1}{2}\sum_{i=1}^{n_{t}} \left[ \left( \left[ \begin{matrix} \tilde{y}_{ijk} \\ \tilde{w}_{ijk} \end{matrix} \right]-\left[ \begin{matrix} \left( \mathbf{X}_{k} \right)_{ij} & 0 \\ 0 & \left( \mathbf{Z}_{k} \right)_{ij} \end{matrix} \right]\mathbf{D}_{jk}\mathbf{u}_{jk} \right)^{T}\mathbf{E}^{-1}\left( \left[ \begin{matrix} \tilde{y}_{ijk} \\ \tilde{w}_{ijk} \end{matrix} \right]-\left[ \begin{matrix} \left( \mathbf{X}_{k} \right)_{ij} & 0 \\ 0 & \left( \mathbf{Z}_{k} \right)_{ij} \end{matrix} \right]\mathbf{D}_{jk}\mathbf{u}_{jk} \right) \right] \right\}\# \\ &\times\left| \Sigma_{jkc} \right|^{-\frac{1}{2}}exp\left\{ -\frac{1}{2}\mathbf{u}_{jk}^{T}{\boldsymbol{\Sigma}_{jk}}^{-1}\mathbf{u}_{jk} \right\} \\ &\propto exp\left\{ -\frac{1}{2}\left( \mathbf{u}_{jk}^{T}\mathbf{C}_{jk}\mathbf{u}_{jk}-2\mathbf{r}_{jk}^{T}\mathbf{u}_{jk} \right) \right\} \\ &\propto MVN\left( \mathbf{C}_{jk}^{-1}\mathbf{r}_{jk},\mathbf{C}_{jk}^{-1} \right)\#(S5) \end{aligned}$$

Where

$$\begin{aligned} \left[ \begin{matrix} \tilde{y}_{ijk} \\ \tilde{w}_{ijk} \end{matrix} \right]&=\left[ \begin{matrix} y_{i} \\ w_{ik} \end{matrix} \right]-\sum_{j=1}^{p_{int}} \left[ \begin{matrix} G_{ij} & 0 \\ 0 & 0 \end{matrix} \right]\left[ \begin{matrix} \gamma_{j} \\ 0 \end{matrix} \right]-\sum_{j^{'}\neq j}^{p_{k}} \left[ \begin{matrix} \left( \mathbf{X}_{k} \right)_{ij^{'}} & 0 \\ 0 & \left( \mathbf{Z}_{k} \right)_{ij^{'}} \end{matrix} \right]\mathbf{D}_{j^{'}k}\mathbf{u}_{j^{'}k}-\sum_{k^{'}\neq k} \left[ \begin{matrix} \left( \mathbf{X}_{k} \right)_{,ij} & 0 \\ 0 & \left( \mathbf{Z}_{k} \right)_{ij} \end{matrix} \right]\mathbf{D}_{jk^{'}}\mathbf{u}_{j^{'}k} \\ \mathbf{C}_{jk}&=\sum_{i=1}^{n_{t}} \mathbf{D}_{jk}^{T}\left[ \begin{matrix} \left( \mathbf{X}_{k} \right)_{ij} & 0 \\ 0 & \left( \mathbf{Z}_{k} \right)_{ij} \end{matrix} \right]\mathbf{E}^{-1}\left[ \begin{matrix} \left( \mathbf{X}_{k} \right)_{ij} & 0 \\ 0 & \left( \mathbf{Z}_{k} \right)_{ij} \end{matrix} \right]\mathbf{D}_{jk}+\boldsymbol{\Sigma}_{jk}^{-1} \\ \mathbf{r}_{jk}&=\sum_{i=1}^{n_{t}} \left[ \begin{matrix} \tilde{y}_{ijk} \\ \tilde{w}_{ijk} \end{matrix} \right]^{T}\mathbf{E}^{-1}\left[ \begin{matrix} \left( \mathbf{X}_{k} \right)_{ij} & 0 \\ 0 & \left( \mathbf{Z}_{k} \right)_{ij} \end{matrix} \right]\mathbf{D}_{jk}\#(S6)\# \end{aligned}$$

Single-site Gibbs sample I is used to estimate maker effects^1^. For convenience, from now on let “1” denotes one trait and “2” denotes the other trait. After dropping factors that do not involve $u_{jk,1}$, we have

$$\begin{aligned} f\left( u_{jk,1}|{\delta_{jk,1}\boldsymbol{,}u}_{jk,2},\pi=c,\mathbf{y}\boldsymbol{,}\mathbf{w}_{k}\boldsymbol{,}\boldsymbol{\Sigma}_{jk} \right)&\propto exp\left\{ -\frac{1}{2}\left[ \left[ \begin{matrix} u_{jk,1} \\ u_{jk,2} \end{matrix} \right]^{T}\left[ \begin{matrix} C_{jk,11} & C_{jk,12} \\ C_{jk,21} & C_{jk,22} \end{matrix} \right]\left[ \begin{matrix} u_{jk,1} \\ u_{jk,2} \end{matrix} \right]-2\left[ \begin{matrix} r_{jk,1} & r_{jk,2} \end{matrix} \right]\left[ \begin{matrix} u_{jk,1} \\ u_{jk,2} \end{matrix} \right] \right] \right\}\# \\ &\propto exp\left\{ -\frac{1}{2}\left[ \left[ \begin{matrix} u_{jk,1} \\ u_{jk,2} \end{matrix} \right]^{T}\left[ \begin{matrix} C_{jk,11} & C_{jk,12} \\ C_{jk,21} & C_{jk,22} \end{matrix} \right]\left[ \begin{matrix} u_{jk,1} \\ u_{jk,2} \end{matrix} \right]-2\left[ \begin{matrix} r_{jk,1} & r_{jk,2} \end{matrix} \right]\left[ \begin{matrix} u_{jk,1} \\ u_{jk,2} \end{matrix} \right] \right] \right\} \\ &\propto N\left( \left( C_{jk,11} \right)^{-1}\left( r_{jk,1}-C_{jk,12}u_{jk,2} \right),\left( C_{jk,11} \right)^{-1} \right) \\ &\propto N\left( \hat{u}_{jk,1},\left( C_{jk,11} \right)^{-1} \right)\#(S7) \end{aligned}$$

Where

$$\begin{aligned} \mathbf{C}_{jk}&=\sum_{i=1}^{n_{t}} \mathbf{D}_{jk}^{T}\left[ \begin{matrix} \left( \mathbf{X}_{k} \right)_{ij} & 0 \\ 0 & \left( \mathbf{Z}_{k} \right)_{ij} \end{matrix} \right]\mathbf{E}^{-1}\left[ \begin{matrix} \left( \mathbf{X}_{k} \right)_{ij} & 0 \\ 0 & \left( \mathbf{Z}_{k} \right)_{ij} \end{matrix} \right]\mathbf{D}_{jk}+\boldsymbol{\Sigma}_{jk}^{-1} \\ &=\sum_{i=1}^{n_{t}} \left[ \begin{matrix} \delta_{jk,1}^{2}\left( \mathbf{X}_{k} \right)_{ij}^{2}\left( \mathbf{E}^{-1} \right)_{11} & \delta_{jk,1}\delta_{jk,2}\left( \mathbf{X}_{k} \right)_{ij}\left( \mathbf{Z}_{k} \right)_{ij}\left( \mathbf{E}^{-1} \right)_{12} \\ \delta_{jk,1}\delta_{jk,2}\left( \mathbf{X}_{k} \right)_{ij}\left( \mathbf{Z}_{k} \right)_{ij}\left( \mathbf{E}^{-1} \right)_{21} & \delta_{jk,2}^{2}\left( \mathbf{Z}_{k} \right)_{ij}^{2}\left( \mathbf{E}^{-1} \right)_{22} \end{matrix} \right]+\left[ \begin{matrix} \left( \boldsymbol{\Sigma}_{jk}^{-1} \right)_{11} & \left( \boldsymbol{\Sigma}_{jk}^{-1} \right)_{12} \\ \left( \boldsymbol{\Sigma}_{jk}^{-1} \right)_{21} & \left( \boldsymbol{\Sigma}_{jk}^{-1} \right)_{22} \end{matrix} \right] \\ &=\left[ \begin{matrix} \delta_{jk,1}^{2}\left( \mathbf{E}^{-1} \right)_{11}\left( \mathbf{X}_{k} \right)_{j}^{T}\left( \mathbf{X}_{k} \right)_{j}+\left( \boldsymbol{\Sigma}_{jk}^{-1} \right)_{11} & \delta_{jk,1}\delta_{jk,2}\left( \mathbf{E}^{-1} \right)_{12}\left( \mathbf{X}_{k} \right)_{j}^{T}\left( \mathbf{Z}_{k} \right)_{j}+\left( \boldsymbol{\Sigma}_{jk}^{-1} \right)_{12} \\ \delta_{jk,1}\delta_{jk,2}\left( \mathbf{E}^{-1} \right)_{21}\left( \mathbf{X}_{k} \right)_{j}^{T}\left( \mathbf{Z}_{k} \right)_{j}+\left( \boldsymbol{\Sigma}_{jk}^{-1} \right)_{21} & \delta_{jk,2}^{2}\left( \mathbf{Z}_{k} \right)_{j}^{T}\left( \mathbf{Z}_{k} \right)_{j}\left( \mathbf{E}^{-1} \right)_{22}+\left( \boldsymbol{\Sigma}_{jk}^{-1} \right)_{22} \end{matrix} \right] \\ \mathbf{r}_{jk}&=\sum_{i=1}^{n_{t}} \left[ \begin{matrix} \tilde{y}_{ijk} \\ \tilde{w}_{ijk} \end{matrix} \right]^{T}\mathbf{E}^{-1}\left[ \begin{matrix} \left( \mathbf{X}_{k} \right)_{ij} & 0 \\ 0 & \left( \mathbf{Z}_{k} \right)_{ij} \end{matrix} \right]\mathbf{D}_{jk} \\ &=\left[ \begin{matrix} \delta_{jk,1}\sum_{i=1}^{n_{t}} {\tilde{y}_{ijk}\left( \mathbf{X}_{k} \right)}_{ij}\left( \mathbf{E}^{-1} \right)_{11} & \delta_{jk,2}\sum_{i=1}^{n_{t}} \tilde{w}_{ijk}\left( \mathbf{Z}_{k} \right)_{ij}\left( \mathbf{E}^{-1} \right)_{22} \end{matrix} \right]\#(S8)\# \end{aligned}$$

Note that when $\delta_{jk,1}=0$,

$$\begin{aligned} \mathbf{C}_{jk}&=\left[ \begin{matrix} C_{jk,11}^{0} & C_{jk,12}^{0} \\ C_{jk,21}^{0} & C_{jk,22}^{0} \end{matrix} \right]=\left[ \begin{matrix} \left( \boldsymbol{\Sigma}_{jk}^{-1} \right)_{11} & \left( \boldsymbol{\Sigma}_{jk}^{-1} \right)_{12} \\ \left( \boldsymbol{\Sigma}_{jk}^{-1} \right)_{21} & \left( \boldsymbol{\Sigma}_{jk}^{-1} \right)_{22}+\delta_{jk,2}^{2}\left( \mathbf{E}^{-1} \right)_{22}\sum_{i=1}^{n_{t}} \left( \mathbf{Z}_{k} \right)_{ij}^{2} \end{matrix} \right] \\ \mathbf{r}_{jk}&=\left[ \begin{matrix} r_{jk,1}^{0} & r_{jk,2}^{0} \end{matrix} \right]=\left[ \begin{matrix} 0 & \delta_{jk,2}\sum_{i=1}^{n_{t}} \tilde{w}_{ijk}\left( \mathbf{Z}_{k} \right)_{ij}\left( \mathbf{E}^{-1} \right)_{22} \end{matrix} \right]\#(S9) \end{aligned}$$

When $\delta_{jk,1}=1$,

$$\begin{aligned} \mathbf{C}_{jk}&=\left[ \begin{matrix} C_{jk,11}^{1} & C_{jk,12}^{1} \\ C_{jk,21}^{1} & C_{jk,22}^{1} \end{matrix} \right]=\left[ \begin{matrix} \delta_{jk,1}^{2}\left( \mathbf{E}^{-1} \right)_{11}\left( \mathbf{X}_{k} \right)_{j}^{T}\left( \mathbf{X}_{k} \right)_{j}+\left( \boldsymbol{\Sigma}_{jk}^{-1} \right)_{11} & \delta_{jk,1}\delta_{jk,2}\left( \mathbf{E}^{-1} \right)_{12}\left( \mathbf{X}_{k} \right)_{j}^{T}\left( \mathbf{Z}_{k} \right)_{j}+\left( \boldsymbol{\Sigma}_{jk}^{-1} \right)_{12} \\ \delta_{jk,1}\delta_{jk,2}\left( \mathbf{E}^{-1} \right)_{21}\left( \mathbf{X}_{k} \right)_{j}^{T}\left( \mathbf{Z}_{k} \right)_{j}+\left( \boldsymbol{\Sigma}_{jk}^{-1} \right)_{21} & \delta_{jk,2}^{2}\left( \mathbf{Z}_{k} \right)_{j}^{T}\left( \mathbf{Z}_{k} \right)_{j}\left( \mathbf{E}^{-1} \right)_{22}+\left( \boldsymbol{\Sigma}_{jk}^{-1} \right)_{22} \end{matrix} \right] \\ \mathbf{r}_{jk}&=\left[ \begin{matrix} r_{jk,1}^{1} & r_{jk,1}^{1} \end{matrix} \right]=\left[ \begin{matrix} \delta_{jk,1}\sum_{i=1}^{n_{t}} {\tilde{y}_{ijk}\left( \mathbf{X}_{k} \right)}_{ij}\left( \mathbf{E}^{-1} \right)_{11} & \delta_{jk,2}\sum_{i=1}^{n_{t}} \tilde{w}_{ijk}\left( \mathbf{Z}_{k} \right)_{ij}\left( \mathbf{E}^{-1} \right)_{22} \end{matrix} \right]\#(S10) \end{aligned}$$

Thus, when $\delta_{jk,1}=0$, the full conditional distribution of $u_{jk,1}$ is

$$\begin{aligned} f\left( u_{jk,1}|{\delta_{jk,1}\boldsymbol{,}u}_{jk,2},\pi=c,\mathbf{y}\boldsymbol{,}\mathbf{w}_{k}\boldsymbol{,}\boldsymbol{\Sigma}_{jk} \right)\propto N\left( \hat{u_{jk,1}^{0}},\left( C_{jk,11}^{0} \right)^{-1} \right)=N\left( {-\left( \boldsymbol{\Sigma}_{jk}^{-1} \right)}_{11}\left( \boldsymbol{\Sigma}_{jk}^{-1} \right)_{12}u_{jk,2},\left( \boldsymbol{\Sigma}_{jk}^{-1} \right)_{11} \right)\#\left( S11 \right) \end{aligned}$$

Thus, when $\delta_{jk,1}=1$, the full conditional distribution of $u_{jk,1}$ is

$$\begin{aligned} f\left( u_{jk,1}|{\delta_{jk,1}\boldsymbol{,}u}_{jk,2},\pi=c,\mathbf{y}\boldsymbol{,}\mathbf{w}_{k}\boldsymbol{,}\boldsymbol{\Sigma}_{jk} \right)\propto N\left( \hat{u_{jk,1}^{1}},\left( C_{jk,11}^{1} \right)^{-1} \right)=N\left( \left( C_{jk,11}^{1} \right)^{-1}\left( r_{jk,1}-C_{jk,12}^{1}u_{jk,2} \right),\left( C_{jk,11}^{1} \right)^{-1} \right)\#\left( S12 \right) \end{aligned}$$

$\delta_{jk,1}$ can be drawn from this categorical distribution by calculating the membership probabilities

$$\begin{aligned} \Pr\left( \delta_{jk,1}=c|\mathbf{y},\mathbf{w}_{k},\mathbf{G},\boldsymbol{\gamma},\mathbf{u}_{jk},\boldsymbol{\Sigma}_{jk},\sigma_{e}^{2},\boldsymbol{\sigma}_{\boldsymbol{\epsilon}}^{2} \right)=\frac{f\left( \mathbf{y},\mathbf{w}_{k}\boldsymbol{|}\delta_{jk,1}=c,\mathbf{G},\boldsymbol{\gamma},\mathbf{u}_{jk},\boldsymbol{\Sigma}_{jk},\sigma_{e}^{2},\boldsymbol{\sigma}_{\boldsymbol{\epsilon}}^{2} \right)f\left( \delta_{jk,1}=c \right)}{\sum_{c^{'}=0}^{1} f\left( \mathbf{y},\mathbf{w}_{k}\boldsymbol{|}\delta_{jk,1}=c^{'},\mathbf{G},\boldsymbol{\gamma},\mathbf{u}_{jk},\boldsymbol{\Sigma}_{jk},\sigma_{e}^{2},\boldsymbol{\sigma}_{\boldsymbol{\epsilon}}^{2} \right)f\left( \delta_{jk,1}=c^{'} \right)},c=0,1\#(S13)\# \end{aligned}$$

Where $f\left( \mathbf{y},\mathbf{w}_{k}|\delta_{jk}=c,\mathbf{G},\boldsymbol{\gamma},\mathbf{u}_{jk},\boldsymbol{\Sigma}_{jk},\delta_{jk},\sigma_{e}^{2},\boldsymbol{\sigma}_{\boldsymbol{\epsilon}}^{2} \right)=\int f\left( \mathbf{y},\mathbf{w}_{k}|\delta_{jk,1}=c,\mathbf{u} \right)f\left( u_{jk,1}|{\delta_{jk,1},u}_{jk,2} \right)du_{jk,1}$. When “1” denotes the complex trait, $f\left( \delta_{jk,1}=1 \right)=\pi_{\beta}$, and when “1” denotes the $k$-th molecular phenotype, $f\left( \delta_{jk,1}=1 \right)=\pi_{\alpha}$.

Assuming the prior of $\boldsymbol{\Sigma}_{jk}$ is inverse Wishart distribution

$$\begin{aligned} p\left( \boldsymbol{\Sigma}_{jk} \right)&=\frac{\left| \mathbf{S} \right|^{\frac{v}{2}}\left| \boldsymbol{\Sigma}_{jk} \right|^{-\frac{\left( v+p+1 \right)}{2}}exp\left\{ -tr\left( \mathbf{S}\boldsymbol{\Sigma}_{jk}^{-1} \right) \right\}}{2^{\frac{vp}{2}}\Gamma\left( \frac{v}{2} \right)}\sim W^{-1}\left( \mathbf{S},v \right)\#(S14)\# \end{aligned}$$

Where $v=4$ and $\mathbf{S}\boldsymbol{=}\left[ \begin{matrix} \frac{h_{snp}^{2}}{n_{t}} & 0 \\ 0 & \frac{h_{cis,k}^{2}}{n_{m}} \end{matrix} \right]$.

Then the full conditional distribution of $\boldsymbol{\Sigma}_{jk}$ using distribution prior is

$$\begin{aligned} f\left( \boldsymbol{\Sigma}_{jk}\boldsymbol{|}\mathbf{u}\} \right)&\propto p\left( \mathbf{u}\boldsymbol{|}\boldsymbol{\Sigma}_{jk} \right)p\left( \boldsymbol{\Sigma}_{jk} \right)\# \\ &\propto\left| \boldsymbol{\Sigma}_{jk} \right|^{-p_{k}/2}exp\left\{ -\frac{1}{2}\sum_{j=1}^{p_{k}} \mathbf{u}^{T}\boldsymbol{\Sigma}_{jk}^{-1}\mathbf{u} \right\}\left| \mathbf{S} \right|^{v/2}\left| \boldsymbol{\Sigma}_{jk} \right|^{-\left( v+p+1 \right)/2}exp\left\{ -tr\left( \mathbf{S}\boldsymbol{\Sigma}_{jk}^{-1} \right) \right\} \\ &\propto\left| \boldsymbol{\Sigma}_{jk} \right|^{-\left[ p_{k}-v+p+1 \right]/2}exp\left\{ -\frac{1}{2}tr\left[ \left( \sum_{j=1}^{p_{k}} \mathbf{u}_{jk}^{T}\mathbf{u}_{jk}\boldsymbol{+}\mathbf{S} \right)\boldsymbol{\Sigma}_{jk}^{-1} \right] \right\}\left\lceil\mathbf{S} \right\rceil^{v/2}\left\lceil\boldsymbol{\Sigma}_{jk} \right\rceil^{-\left( v+p+1 \right)/2}exp\left\{ -tr\left( \mathbf{S}\boldsymbol{\Sigma}_{jk}^{-1} \right) \right\} \\ &\propto W^{-1}\left( \sum_{j=1}^{p_{k}} \mathbf{u}_{jk}^{T}\mathbf{u}_{jk}\boldsymbol{+}\mathbf{S},v+p_{k} \right)\#(S15) \end{aligned}$$

Where $p=2$ since $\boldsymbol{\Sigma}_{jk}$ is always $2\times2$ matrix for the $k$-th molecular phenotype.

The full conditional distribution of $\sigma_{e}^{2}$ is

$$\begin{aligned} f\left( \sigma_{e}^{2}|\mathbf{y},\boldsymbol{\gamma},\boldsymbol{\beta,}\mathbf{G,}\boldsymbol{\gamma}\boldsymbol{,}\sigma_{e}^{2} \right)&\propto f\left( \mathbf{y}|\boldsymbol{\gamma},\boldsymbol{\beta,}\mathbf{G,}\boldsymbol{\gamma}\boldsymbol{,}\sigma_{e}^{2} \right)f\left( \sigma_{e}^{2} \right) \\ &\propto\left( \sigma_{e}^{2} \right)^{-\frac{n_{g}}{2}} exp\left\{ -\frac{\left( \mathbf{y}\boldsymbol{-}\mathbf{G}\boldsymbol{\gamma-}\boldsymbol{X\beta} \right)^{T}\left( \mathbf{y}\boldsymbol{-}\mathbf{G}\boldsymbol{\gamma-}\boldsymbol{X\beta} \right)}{2\sigma_{e}^{2}} \right\}\left( \sigma_{e}^{2} \right)^{-\frac{\nu_{e}+2}{2}}exp\left\{ -\frac{\nu_{e}S_{e}^{2}}{2\sigma_{e}^{2}} \right\} \\ &\propto\left( \sigma_{e}^{2} \right)^{-\frac{n_{g}+\nu_{e}+2}{2}} exp\left\{ -\frac{\mathbf{e}^{T}\mathbf{e}+\nu_{e}S_{e}^{2}}{2\sigma_{e}^{2}} \right\} \\ &=\chi^{-2}\left( \hat{\nu}_{e},\hat{S}_{e}^{2} \right)\#(S16)\# \end{aligned}$$

Where $\tilde{\nu}_{e}=n_{g}+\nu_{e}$ and $\tilde{S}_{e}^{2}=\left( \mathbf{e}^{T}\mathbf{e}+\nu_{e}S_{e}^{2} \right)/{\tilde{\nu}_{e}}$.

The full conditional distribution of $\sigma_{\boldsymbol{\epsilon}_{k}}^{2}$ is

$$\begin{aligned} f\left( \sigma_{\epsilon_{k}}^{2}|\mathbf{w}_{k},\boldsymbol{\alpha}_{k},\sigma_{\epsilon_{k}}^{2} \right)&\propto f\left( \mathbf{w}_{k}|\boldsymbol{\alpha}_{k},\sigma_{\epsilon_{k}}^{2} \right)f\left( \sigma_{\epsilon_{k}}^{2} \right) \\ &\propto\left( \sigma_{\boldsymbol{\epsilon}_{k}}^{2} \right)^{-\frac{n_{mo}}{2}} exp\left\{ -\frac{\left( \mathbf{w}_{k}-\mathbf{X}_{k}\boldsymbol{\alpha}_{k} \right)^{T}\left( \mathbf{w}_{k}-\mathbf{X}_{k}\boldsymbol{\alpha}_{k} \right)}{2\sigma_{\boldsymbol{\epsilon}_{k}}^{2}} \right\}\left( \sigma_{\epsilon_{k}}^{2} \right)^{-\frac{\nu_{e}+2}{2}}exp\left\{ -\frac{\nu_{e}S_{e}^{2}}{2\sigma_{\epsilon_{k}}^{2}} \right\} \\ &\propto\left( \sigma_{\boldsymbol{\epsilon}_{k}}^{2} \right)^{-\frac{n_{mo}+\nu_{\boldsymbol{\epsilon}_{k}}+2}{2}} exp\left\{ -\frac{\boldsymbol{\epsilon}_{k}^{T}\boldsymbol{\epsilon}_{k}+\nu_{\boldsymbol{\epsilon}_{k}}S_{\boldsymbol{\epsilon}_{k}}^{2}}{2\sigma_{\boldsymbol{\epsilon}_{k}}^{2}} \right\} \\ &=\chi^{-2}\left( \hat{\nu}_{e},\hat{S}_{e}^{2} \right)\#(S17)\# \end{aligned}$$

Where $\tilde{\nu}_{\boldsymbol{\epsilon}_{k}}=n_{k}+\nu_{\boldsymbol{\epsilon}_{k}}$ and $\tilde{S}_{\boldsymbol{\epsilon}_{k}}^{2}=\left( \boldsymbol{\epsilon}^{T}\boldsymbol{\epsilon}_{k}+\nu_{\boldsymbol{\epsilon}_{k}}S_{\boldsymbol{\epsilon}_{k}}^{2} \right)/{\tilde{\nu}_{\boldsymbol{\epsilon}_{k}}}$.

**Adjust residual variance in the low-rank model.** Our low-rank model is derived from the summary-level model with a full-LD matrix and needs to be adjusted when calculating the sum of squared errors (SSE). In the summary-level BayesC model^2,3^, when the $j$-th marker effect $\beta_{j}$ is sampled, take GWAS as an example, its full conditional distribution can be constructed as $N(\frac{r_{j}}{l_{jc}},\frac{\sigma_{e}^{2}}{l_{jc}})$, where $l_{jc}=\mathbf{X}_{j}^{T}\mathbf{X}_{j}+{\sigma_{e}^{2}}/{\sigma_{\beta}^{2}}$, $r_{j}=\mathbf{X}_{j}^{T}\mathbf{w}$, and $\mathbf{w}=\mathbf{y}\boldsymbol{-}\mathbf{X}_{\boldsymbol{-}j}\boldsymbol{\beta}_{\boldsymbol{-}j}$, SSE in the model can be set as

$$\begin{aligned} SSE&=\left( \mathbf{y}\boldsymbol{-}\mathbf{X}\boldsymbol{\beta} \right)^{T}\left( \mathbf{y}\boldsymbol{-}\mathbf{X}\boldsymbol{\beta} \right) \\ &=\mathbf{y}^{T}\mathbf{y}\boldsymbol{-}{\boldsymbol{\beta}^{T}\mathbf{X}}^{T}\mathbf{y}\boldsymbol{-}{\boldsymbol{\beta}^{T}\mathbf{X}}^{T}\mathbf{y}\boldsymbol{+}{\boldsymbol{\beta}^{T}\mathbf{X}}^{T}\mathbf{X}\boldsymbol{\beta} \\ &=\mathbf{y}^{\mathbf{T}}\mathbf{y-}\boldsymbol{\beta}^{T}\mathbf{Db-}\boldsymbol{\beta}^{T}\mathbf{Db+}{\boldsymbol{\beta}^{T}\mathbf{X}}^{T}\mathbf{X}\boldsymbol{\beta} \\ &=\mathbf{y}^{\mathbf{T}}\mathbf{y-}\boldsymbol{\beta}^{T}\mathbf{Db-}\boldsymbol{\beta}^{T}\mathbf{r}^{\mathbf{*}} \\ &=\mathbf{y}^{\mathbf{T}}\mathbf{y-}\boldsymbol{\beta}^{T}\left( \mathbf{Db+}\mathbf{r}^{\mathbf{*}} \right)\#(S18)\# \end{aligned}$$

Where $\mathbf{r}^{\mathbf{*}}\mathbf{=}\mathbf{X}^{T}\mathbf{y-}\mathbf{X}^{T}\mathbf{X}\boldsymbol{\beta}$ is corrected right-hand side in the RHS updating scheme, where $\mathbf{X}^{T}\mathbf{X=}\mathbf{R}/{n_{g}}$, $\mathbf{X}^{T}\mathbf{y= Db}$ based on the least squares solutions.

Similarly, SSE in the low-rank summary model is

$$\begin{aligned} \mathbf{SSE&=}\mathbf{e}^{T}\mathbf{e=}\left( \mathbf{y-X}\boldsymbol{\beta} \right)^{T}\left( \mathbf{y-X}\boldsymbol{\beta} \right) \\ \mathbf{&=}\mathbf{y}^{T}\mathbf{y-}\boldsymbol{\beta}^{T}\mathbf{Db-}\boldsymbol{\beta}^{T}\mathbf{Db+}{n_{g}\boldsymbol{\beta}}^{T}\mathbf{R}_{\mathbf{X}}\boldsymbol{\beta} \\ \mathbf{&=}\mathbf{y}^{T}\mathbf{y-}\boldsymbol{\beta}^{T}\left( n_{g}\mathbf{I} \right)\mathbf{b-}\boldsymbol{\beta}^{T}\left( \mathbf{U}_{\mathbf{X}}\boldsymbol{\Lambda}_{\mathbf{X}}^{\frac{\mathbf{1}}{\mathbf{2}}}\boldsymbol{\cdot}\boldsymbol{\Lambda}_{\mathbf{X}}^{-\frac{1}{2}}\mathbf{U}_{\mathbf{X}}^{T} \right)\left( n_{g}\mathbf{I} \right)\mathbf{b+}n_{g}\left( \mathbf{U}_{\mathbf{X}}\boldsymbol{\Lambda}_{\mathbf{X}}^{\frac{\mathbf{1}}{\mathbf{2}}}\boldsymbol{\cdot}\boldsymbol{\Lambda}_{\mathbf{X}}^{\frac{\mathbf{1}}{\mathbf{2}}}\mathbf{U}_{\mathbf{X}}^{T} \right)\boldsymbol{\beta} \\ \mathbf{&=}\mathbf{y}^{T}\mathbf{y-}\boldsymbol{\beta}^{T}\left( n_{g}\mathbf{I} \right)\mathbf{b-}\boldsymbol{\beta}^{T}\left[ n_{g}\mathbf{Q}_{\mathbf{X}}^{T}\left( \boldsymbol{\Lambda}_{\mathbf{X}}^{-\frac{1}{2}}\mathbf{U}_{\mathbf{X}}^{T}\mathbf{b-}\mathbf{Q}_{\mathbf{X}}\boldsymbol{\beta} \right) \right] \\ \mathbf{&=}\mathbf{y}^{T}\mathbf{y-}\boldsymbol{\beta}^{T}n_{g}\mathbf{Ib-}{n_{g}\boldsymbol{\beta}}^{T}\mathbf{Q}_{\mathbf{X}}^{T}{\hat{\mathbf{r}}}^{\mathbf{*}} \\ \mathbf{&=}n_{g}\left[ \frac{\mathbf{y}^{T}\mathbf{y}}{n_{g}}\mathbf{-}\boldsymbol{\beta}^{T}\left( \mathbf{b+}\mathbf{Q}_{\mathbf{X}}^{T}{\hat{\mathbf{r}}}^{\mathbf{*}} \right) \right]\boldsymbol{\#(}S19\boldsymbol{)\#\#\#} \end{aligned}$$

Where ${\hat{\mathbf{r}}}^{\mathbf{*}}\boldsymbol{=}\boldsymbol{\Lambda}_{\mathbf{X}}^{-\frac{1}{2}}\mathbf{U}_{\mathbf{X}}^{T}\mathbf{b-}\mathbf{Q}_{\mathbf{X}}\boldsymbol{\beta}$ and $\mathbf{Q}_{\mathbf{X}}\mathbf{=}\boldsymbol{\Lambda}_{\mathbf{X}}^{\frac{1}{2}}\mathbf{U}_{\mathbf{X}}^{T}$.

**MCMC sampling in the low-rank summary-level model.** The joint distribution of data and all parameters in the low-rank summary-level model is

$$\begin{aligned} f\left( \hat{\mathbf{b}},\hat{\mathbf{a}},\boldsymbol{\beta},\boldsymbol{\alpha},\boldsymbol{\Sigma},\boldsymbol{\pi},\sigma_{e}^{2},\boldsymbol{\sigma}_{\boldsymbol{\epsilon}}^{2} \right)&\propto\prod_{k=1}^{K} exp\left\{ -\frac{1}{2}\sum_{l} \left[ \mathbf{A}^{T}\mathbf{E}^{-1}\mathbf{A} \right] \right\}\# \\ &\times\prod_{k=1}^{K} \prod_{j=1}^{p_{k}} \left| \boldsymbol{\Sigma}_{jk} \right|^{-\frac{1}{2}}exp\left\{ -\frac{1}{2}\mathbf{u}_{jk}^{T}{\boldsymbol{\Sigma}_{jk}}^{-1}\mathbf{u}_{jk} \right\} \\ &\times\prod_{k=1}^{K} \frac{exp\left( {-\nu_{\epsilon,k}S_{\epsilon,k}^{2}}/{2\left( \sigma_{\epsilon_{a_{k}}^{*}}^{2} \right)} \right)}{\left( \sigma_{\epsilon_{a_{k}}^{*}}^{2} \right)^{1+{\nu_{\epsilon,k}}/2}} \\ &\times\prod_{k=1}^{K} \frac{\left| \mathbf{S}_{\Sigma} \right|^{v_{\Sigma}/2}\left| \boldsymbol{\Sigma}_{k} \right|^{-\left( v_{\Sigma}+2+1 \right)/2}exp\left\{ -tr\left( \mathbf{S}_{\Sigma}\boldsymbol{\Sigma}_{k}^{-1} \right) \right\}}{2^{{2v_{\Sigma}}/2}\Gamma\left( {v_{\Sigma}}/2 \right)} \\ &\times\prod_{j=1}^{p_{int}} \left( 2{\pi\sigma}_{\gamma}^{2} \right)^{-\frac{1}{2}}exp\left[ \frac{\gamma_{j}^{2}}{2\sigma_{\gamma}^{2}} \right] \\ &\times\frac{\exp\left( {-\nu_{\gamma}S_{\gamma}^{2}}/{2\sigma_{\gamma}^{2}} \right)}{\left( \sigma_{\gamma}^{2} \right)^{1+{\nu_{\gamma}}/2}} \\ &\times\frac{\exp\left( {-\nu_{e_{\beta}^{*}}S_{e_{\beta}^{*}}^{2}}/{2\sigma_{e_{\beta}^{*}}^{2}} \right)}{\left( \sigma_{e_{\beta}^{*}}^{2} \right)^{1+{\nu_{e_{\beta}^{*}}}/2}}\#(S20) \end{aligned}$$

Where $\mathbf{E}=\left[ \begin{matrix} \sigma_{e_{\beta}^{*}}^{2} & 0 \\ 0 & \sigma_{\epsilon_{a_{k}}^{*}}^{2} \end{matrix} \right]$, $\mathbf{A}=\left[ \begin{matrix} {\hat{\mathbf{b}}}_{l} \\ {\hat{\mathbf{a}}}_{lk} \end{matrix} \right]-\sum_{j=1}^{p_{int}} \left[ \begin{matrix} \left( \mathbf{Q}_{\mathbf{G}} \right)_{lj} & 0 \\ 0 & 0 \end{matrix} \right]\left[ \begin{matrix} \gamma_{j} \\ 0 \end{matrix} \right]-\sum_{j=1}^{p_{k}} \left[ \begin{matrix} \left( \mathbf{Q}_{\mathbf{X},k} \right)_{lj} & 0 \\ 0 & \left( \mathbf{Q}_{\mathbf{Z},k} \right)_{lj} \end{matrix} \right]\mathbf{D}_{jk}\mathbf{u}_{jk}$.

The full conditional distribution of $\gamma_{j}$ is

$$\begin{aligned} f\left( \gamma_{j}|\hat{\boldsymbol{b}},\mathbf{Q}_{\mathbf{X}}\boldsymbol{,}\boldsymbol{\beta,} \boldsymbol{\gamma}_{-j}\eta_{j}=1,\sigma_{e}^{2} \right)&\propto exp\left\{ -\frac{1}{2\sigma_{e}^{2}}\left( \tilde{\mathbf{b}}-\left( \mathbf{Q}_{\mathbf{G}} \right)_{j}\gamma_{j} \right)^{T}\left( \tilde{\mathbf{b}}-\left( \mathbf{Q}_{\mathbf{G}} \right)_{j}\gamma_{j} \right) \right\}exp\left\{ -\frac{\gamma_{j}^{2}}{2\sigma_{\gamma}^{2}} \right\}\# \\ &\propto N\left( \frac{r_{j}}{C_{j}},\frac{\sigma_{e}^{2}}{C_{j}} \right)\#(S21) \end{aligned}$$

Where

$$\begin{aligned} \tilde{\mathbf{b}}\mathbf{&=}\hat{\boldsymbol{b}}-\sum_{k} \left( \mathbf{Q}_{\mathbf{X}} \right)_{k}\boldsymbol{\beta}_{k}\mathbf{-}\sum_{j^{'}\neq j} \left( \mathbf{Q}_{\mathbf{G}} \right)_{j^{'}}\gamma_{j^{'}}\# \\ r_{j}&=\left( \mathbf{Q}_{\mathbf{G}} \right)_{j}^{T}\tilde{\mathbf{b}} \\ C_{j}&=\left( \mathbf{Q}_{\mathbf{G}} \right)_{j}^{T}\left( \mathbf{Q}_{\mathbf{G}} \right)_{j}\boldsymbol{+}\frac{\sigma_{e}^{2}}{\sigma_{\gamma}^{2}}\boldsymbol{\#(}S22\boldsymbol{)} \end{aligned}$$

When $\eta_{j}=0$, $\gamma_{j}=0.$

The full conditional distribution $\eta_{j}$ is

$$\begin{aligned} \Pr\left( \eta_{j}=c|\hat{\mathbf{b}},\mathbf{Q}_{\mathbf{X}},\boldsymbol{\beta},\mathbf{Q}_{\mathbf{G}},\boldsymbol{\gamma},\sigma_{\gamma}^{2},\sigma_{e_{\beta}^{*}}^{2} \right)=\frac{f\left( \hat{\mathbf{b}}\boldsymbol{|}\eta_{j}=c,\mathbf{Q}_{\mathbf{X}},\boldsymbol{\beta},\mathbf{Q}_{\mathbf{G}},\boldsymbol{\gamma},\sigma_{\gamma}^{2},\sigma_{e_{\beta}^{*}}^{2} \right)f\left( \eta_{j}=c \right)}{\sum_{c^{'}=0}^{1} f\left( \hat{\mathbf{b}}\boldsymbol{|}\eta_{j}=c^{'},\mathbf{Q}_{\mathbf{X}},\boldsymbol{\beta},\mathbf{Q}_{\mathbf{G}},\boldsymbol{\gamma},\sigma_{\gamma}^{2},\sigma_{e_{\beta}^{*}}^{2} \right)f\left( \eta_{j}=c^{'} \right)},c=0,1\#\#(S23)\# \end{aligned}$$

Where $f\left( \hat{\mathbf{b}}\boldsymbol{|}\eta_{j}=c,\mathbf{Q}_{\mathbf{X}},\boldsymbol{\beta},\mathbf{Q}_{\mathbf{G}},\boldsymbol{\gamma},\sigma_{\gamma}^{2},\sigma_{e_{\beta}^{*}}^{2} \right)=\int f\left( \hat{\mathbf{b}}\boldsymbol{|}\eta_{j}=c,\gamma_{j} \right)f\left( \gamma_{j}|\eta_{j}=c,\sigma_{\gamma}^{2} \right)d\gamma_{j}$ above and $f\left( \eta_{j}=1 \right)=\pi_{\gamma}$.

The full conditional distribution of $\mathbf{u}_{jk}$ is

$$\begin{aligned} f\left( \mathbf{u}_{jk}\boldsymbol{|}\hat{\mathbf{b}},{\hat{\mathbf{a}}}_{k}\mathbf{Q}_{\mathbf{G}}\boldsymbol{,\gamma,}\mathbf{u}_{-jk},\boldsymbol{\Sigma},\delta_{jk},\sigma_{e}^{2},\boldsymbol{\sigma}_{\boldsymbol{\epsilon}}^{2} \right)&\propto exp\left\{ -\frac{1}{2}\sum_{l=1} \left[ \left( \left[ \begin{matrix} \tilde{b}_{ljk} \\ \tilde{a}_{ljk} \end{matrix} \right]-\left[ \begin{matrix} \left( \mathbf{Q}_{\mathbf{X},k} \right)_{lj} & 0 \\ 0 & \left( \mathbf{Q}_{\mathbf{Z},k} \right)_{lj} \end{matrix} \right]\mathbf{D}_{j}\mathbf{u}_{jk} \right)^{T}\mathbf{E}^{-1}\left( \left[ \begin{matrix} \tilde{b}_{ljk} \\ \tilde{a}_{ljk} \end{matrix} \right]-\left[ \begin{matrix} \left( \mathbf{Q}_{\mathbf{X},k} \right)_{lj} & 0 \\ 0 & \left( \mathbf{Q}_{\mathbf{Z},k} \right)_{lj} \end{matrix} \right]\mathbf{D}_{j}\mathbf{u}_{jk} \right) \right] \right\}\# \\ &\times\left| \Sigma_{jkc} \right|^{-\frac{1}{2}}exp\left\{ -\frac{1}{2}\mathbf{u}_{jk}^{T}{\boldsymbol{\Sigma}_{jkc}}^{-1}\mathbf{u}_{jk} \right\} \\ &\propto exp\left\{ -\frac{1}{2}\left( \mathbf{u}_{jk}^{T}\mathbf{C}_{jk}\mathbf{u}_{jk}-2\mathbf{r}_{jk}^{T}\mathbf{u}_{jk} \right) \right\} \\ &\propto MVN\left( \mathbf{C}_{jk}^{-1}\mathbf{r}_{jk},\mathbf{C}_{jk}^{-1} \right)\#(S24) \end{aligned}$$

Where

$$\begin{aligned} \left[ \begin{matrix} \tilde{b}_{ljk} \\ \tilde{a}_{ljk} \end{matrix} \right]&=\left[ \begin{matrix} \hat{b}_{lk} \\ \hat{a}_{lk} \end{matrix} \right]-\sum_{j=1}^{p_{int}} \left[ \begin{matrix} \left( \mathbf{Q}_{\mathbf{G}} \right)_{lj} & 0 \\ 0 & 0 \end{matrix} \right]\left[ \begin{matrix} \gamma_{j} \\ 0 \end{matrix} \right]-\sum_{j^{'}\neq j}^{p_{k}} \left[ \begin{matrix} \left( \mathbf{Q}_{\mathbf{X},k} \right)_{lj^{'}} & 0 \\ 0 & \left( \mathbf{Q}_{\mathbf{Z},k} \right)_{lj^{'}} \end{matrix} \right]\mathbf{D}_{j^{'}k}^{T}\mathbf{u}_{j^{'}k}^{T}-\sum_{k^{'}\neq k} \left[ \begin{matrix} \left( \mathbf{Q}_{\mathbf{X},k^{'}} \right)_{lj} & 0 \\ 0 & \left( \mathbf{Q}_{\mathbf{Z},k^{'}} \right)_{lj} \end{matrix} \right]\mathbf{D}_{jk^{'}}^{T}\mathbf{u}_{jk^{'}}^{T} \\ \mathbf{C}_{jk}&=\sum_{l} \mathbf{D}_{jk}^{T}\left[ \begin{matrix} \left( \mathbf{Q}_{\mathbf{X},k} \right)_{lj} & 0 \\ 0 & \left( \mathbf{Q}_{\mathbf{Z},k} \right)_{lj} \end{matrix} \right]\mathbf{E}^{-1}\left[ \begin{matrix} \left( \mathbf{Q}_{\mathbf{X},k} \right)_{lj} & 0 \\ 0 & \left( \mathbf{Q}_{\mathbf{Z},k} \right)_{lj} \end{matrix} \right]\mathbf{D}_{jk}+\boldsymbol{\Sigma}_{jk}^{-1} \\ \mathbf{r}_{jk}&=\sum_{l} \left[ \begin{matrix} \tilde{b}_{ljk} \\ \tilde{a}_{ljk} \end{matrix} \right]^{T}\mathbf{E}^{-1}\left[ \begin{matrix} \left( \mathbf{Q}_{\mathbf{X},k} \right)_{lj} & 0 \\ 0 & \left( \mathbf{Q}_{\mathbf{Z},k} \right)_{lj} \end{matrix} \right]\mathbf{D}_{jk}\#(S25)\# \end{aligned}$$

Like above individual-level model, when using single-site Gibbs sample I, we can drop factors that do not involve $u_{jk,1}$

$$\begin{aligned} f\left( u_{jk,1}|{\delta_{jk,1}\boldsymbol{,}u}_{jk,2},\hat{\mathbf{b}},{\hat{\mathbf{a}}}_{k}\boldsymbol{,}\mathbf{Q}_{\mathbf{G}}\mathbf{,}\boldsymbol{\gamma}\boldsymbol{,\Sigma},\delta_{jk},\sigma_{e}^{2},\boldsymbol{\sigma}_{\boldsymbol{\epsilon}}^{2} \right)&\propto N\left( \left( C_{jk,11} \right)^{-1}\left( r_{jk,1}-C_{jk,12}u_{jk,2} \right),\left( C_{jk,11} \right)^{-1} \right)\# \\ &\propto N\left( \hat{u}_{jk,1},\left( C_{jk,11} \right)^{-1} \right)\#(S26) \end{aligned}$$

Where,

$$\begin{aligned} \mathbf{C}_{jk}&=\sum_{l} \mathbf{D}_{jk}^{T}\left[ \begin{matrix} \left( \mathbf{Q}_{\mathbf{X},k} \right)_{lj} & 0 \\ 0 & \left( \mathbf{Q}_{\mathbf{Z},k} \right)_{lj} \end{matrix} \right]\mathbf{E}^{-1}\left[ \begin{matrix} \left( \mathbf{Q}_{\mathbf{X},k} \right)_{lj} & 0 \\ 0 & \left( \mathbf{Q}_{\mathbf{Z},k} \right)_{lj} \end{matrix} \right]\mathbf{D}_{jk}+\boldsymbol{\Sigma}_{jk}^{-1} \\ &=\sum_{l} \left[ \begin{matrix} \delta_{jk,1}^{2}\left( \mathbf{Q}_{\mathbf{X,}k} \right)_{lj}^{2}\left( \mathbf{E}^{-1} \right)_{11} & \delta_{jk,1}\delta_{jk,2}\left( \mathbf{Q}_{\mathbf{X,}k} \right)_{lj}\left( \mathbf{Q}_{\mathbf{Z},k} \right)_{lj}\left( \mathbf{E}^{-1} \right)_{12} \\ \delta_{jk,1}\delta_{jk,2}\left( \mathbf{Q}_{\mathbf{X,}k} \right)_{lj}\left( \mathbf{Q}_{\mathbf{Z},k} \right)_{lj}\left( \mathbf{E}^{-1} \right)_{21} & \delta_{jk,2}^{2}\left( \mathbf{Q}_{\mathbf{Z},k} \right)_{lj}^{2}\left( \mathbf{E}^{-1} \right)_{22} \end{matrix} \right]+\left[ \begin{matrix} \left( \boldsymbol{\Sigma}_{jk}^{-1} \right)_{11} & \left( \boldsymbol{\Sigma}_{jk}^{-1} \right)_{12} \\ \left( \boldsymbol{\Sigma}_{jk}^{-1} \right)_{21} & \left( \boldsymbol{\Sigma}_{jk}^{-1} \right)_{22} \end{matrix} \right] \\ &=\left[ \begin{matrix} \delta_{jk,1}^{2}\left( \mathbf{E}^{-1} \right)_{11}\left( \mathbf{Q}_{\mathbf{X,}k} \right)_{j}^{T}\left( \mathbf{Q}_{\mathbf{X,}k} \right)_{j}+\left( \boldsymbol{\Sigma}_{jk}^{-1} \right)_{11} & \delta_{jk,1}\delta_{jk,2}\left( \mathbf{E}^{-1} \right)_{12}\left( \mathbf{Q}_{\mathbf{X,}k} \right)_{j}^{T}\left( \mathbf{Q}_{\mathbf{Z},k} \right)_{j}+\left( \boldsymbol{\Sigma}_{jk}^{-1} \right)_{12} \\ \delta_{jk,1}\delta_{jk,2}\left( \mathbf{E}^{-1} \right)_{21}\left( \mathbf{Q}_{\mathbf{X,}k} \right)_{j}^{T}\left( \mathbf{Q}_{\mathbf{Z},k} \right)_{j}+\left( \boldsymbol{\Sigma}_{jk}^{-1} \right)_{21} & \delta_{jk,2}^{2}\left( \mathbf{Q}_{\mathbf{Z},k} \right)_{j}^{T}\left( \mathbf{Q}_{\mathbf{Z},k} \right)_{j}\left( \mathbf{E}^{-1} \right)_{22}+\left( \boldsymbol{\Sigma}_{jk}^{-1} \right)_{22} \end{matrix} \right] \\ \mathbf{r}_{jk}&=\sum_{l} \left[ \begin{matrix} \tilde{b}_{ljk} \\ \tilde{a}_{ljk} \end{matrix} \right]^{T}\mathbf{E}^{-1}\left[ \begin{matrix} \left( \mathbf{Q}_{\mathbf{X,}k} \right)_{lj} & 0 \\ 0 & \left( \mathbf{Q}_{\mathbf{Z},k} \right)_{lj} \end{matrix} \right]\mathbf{D}_{jk} \\ &=\left[ \begin{matrix} \delta_{jk,1}\left( \mathbf{E}^{-1} \right)_{11}\sum_{l} \tilde{b}_{ljk}\left( \mathbf{Q}_{\mathbf{X,}k} \right)_{lj} & \delta_{jk,2}\left( \mathbf{E}^{-1} \right)_{22}\sum_{l} \tilde{a}_{ljk}\left( \mathbf{Q}_{\mathbf{Z},k} \right)_{lj} \end{matrix} \right]\#\#(S27)\# \end{aligned}$$

Note that when $\delta_{jk,1}=0$,

$$\begin{aligned} \mathbf{C}_{jk}&=\left[ \begin{matrix} C_{jk,11}^{0} & C_{jk,12}^{0} \\ C_{jk,21}^{0} & C_{jk,22}^{0} \end{matrix} \right]=\left[ \begin{matrix} \left( \boldsymbol{\Sigma}_{jk}^{-1} \right)_{11} & \left( \boldsymbol{\Sigma}_{jk}^{-1} \right)_{12} \\ \left( \boldsymbol{\Sigma}_{jk}^{-1} \right)_{21} & \left( \boldsymbol{\Sigma}_{jk}^{-1} \right)_{22}+\delta_{jk,2}^{2}\left( \mathbf{Q}_{\mathbf{Z},k} \right)_{j}^{T}\left( \mathbf{Q}_{\mathbf{Z},k} \right)_{j}\left( \mathbf{E}^{-1} \right)_{22}+\left( \boldsymbol{\Sigma}_{jk}^{-1} \right)_{22} \end{matrix} \right] \\ \mathbf{r}_{jk}&=\left[ \begin{matrix} r_{jk,1}^{0} & r_{jk,2}^{0} \end{matrix} \right]=\left[ \begin{matrix} 0 & \delta_{jk,2}\left( \mathbf{E}^{-1} \right)_{22}\sum_{l} \tilde{a}_{ljk}\left( \mathbf{Q}_{\mathbf{Z},k} \right)_{lj} \end{matrix} \right]\#(S28) \end{aligned}$$

When $\delta_{jk,1}=1$,

$$\begin{aligned} \mathbf{C}_{jk}&=\left[ \begin{matrix} C_{jk,11}^{1} & C_{jk,12}^{1} \\ C_{jk,21}^{1} & C_{jk,22}^{1} \end{matrix} \right]=\left[ \begin{matrix} \delta_{jk,1}^{2}\left( \mathbf{E}^{-1} \right)_{11}\left( \mathbf{Q}_{\mathbf{X},k} \right)_{j}^{T}\left( \mathbf{Q}_{\mathbf{X},k} \right)_{j}+\left( \boldsymbol{\Sigma}_{jk}^{-1} \right)_{11} & \delta_{jk,1}\delta_{jk,2}\left( \mathbf{E}^{-1} \right)_{12}\left( \mathbf{Q}_{\mathbf{Z},k} \right)_{j}^{T}\left( \mathbf{Q}_{\mathbf{Z},k} \right)_{j}+\left( \boldsymbol{\Sigma}_{jk}^{-1} \right)_{12} \\ \delta_{jk,1}\delta_{jk,2}\left( \mathbf{E}^{-1} \right)_{21}\left( \mathbf{Q}_{\mathbf{X},k} \right)_{j}^{T}\left( \mathbf{Q}_{\mathbf{Z},k} \right)_{j}+\left( \boldsymbol{\Sigma}_{jk}^{-1} \right)_{21} & \delta_{jk,2}^{2}\left( \mathbf{Q}_{\mathbf{Z},k} \right)_{j}^{T}\left( \mathbf{Q}_{\mathbf{Z},k} \right)_{j}\left( \mathbf{E}^{-1} \right)_{22}+\left( \boldsymbol{\Sigma}_{jk}^{-1} \right)_{22} \end{matrix} \right] \\ \mathbf{r}_{jk}&=\left[ \begin{matrix} r_{jk,1}^{1} & r_{jk,1}^{1} \end{matrix} \right]=\left[ \begin{matrix} \delta_{jk,1}\left( \mathbf{E}^{-1} \right)_{11}\sum_{l} \tilde{b}_{ljk}\left( \mathbf{Q}_{\mathbf{X},k} \right)_{lj} & \delta_{jk,2}\left( \mathbf{E}^{-1} \right)_{22}\sum_{l} \tilde{a}_{ljk}\left( \mathbf{Q}_{\mathbf{Z},k} \right)_{lj} \end{matrix} \right]\#(S29) \end{aligned}$$

Thus, when $\delta_{jk,1}=0$, the full conditional distribution of $u_{jk,1}$ is

$$\begin{aligned} f\left( u_{jk,1}|{\delta_{jk,1}\boldsymbol{,}u}_{jk,2}\hat{\mathbf{b}},{\hat{\mathbf{a}}}_{k}\boldsymbol{,}\boldsymbol{\Sigma}_{jk} \right)\propto N\left( \hat{u_{jk,1}^{0}},\left( C_{jk,11}^{0} \right)^{-1} \right)=N\left( {-\left( \boldsymbol{\Sigma}_{jk}^{-1} \right)}_{11}\left( \boldsymbol{\Sigma}_{jk}^{-1} \right)_{12}u_{jk,2},\left( \boldsymbol{\Sigma}_{jk}^{-1} \right)_{11} \right)\#\left( S30 \right) \end{aligned}$$

Thus, when $\delta_{jk,1}=1$, the full conditional distribution of $u_{jk,1}$ is

$$\begin{aligned} f\left( u_{jk,1}|{\delta_{jk,1}\boldsymbol{,}u}_{jk,2},\hat{\mathbf{b}},{\hat{\mathbf{a}}}_{k}\boldsymbol{,}\boldsymbol{\Sigma}_{jk} \right)\propto N\left( \hat{u_{jk,1}^{1}},\left( C_{jk,11}^{1} \right)^{-1} \right)=N\left( \left( C_{jk,11}^{1} \right)^{-1}\left( r_{jk,1}-C_{jk,12}^{1}u_{jk,2} \right),\left( C_{jk,11}^{1} \right)^{-1} \right)\#\left( S31 \right) \end{aligned}$$

$\delta_{jk,1}$ can be drawn from this categorical distribution by calculating the membership probabilities

$$\begin{aligned} \Pr\left( \delta_{jk,1}=c|\hat{\mathbf{b}},{\hat{\mathbf{a}}}_{k},\mathbf{u}_{jk},\boldsymbol{\Sigma}_{jk},\delta_{jk},\sigma_{e_{\beta}^{*}}^{2},\sigma_{\epsilon_{a_{k}}^{*}}^{2} \right)=\frac{f\left( \hat{\mathbf{b}},{\hat{\mathbf{a}}}_{k}\boldsymbol{|}\delta_{jk,1}=c,\mathbf{u}_{jk}\boldsymbol{,}\boldsymbol{\Sigma}_{jk}\boldsymbol{,}\sigma_{e_{\beta}^{*}}^{2},\sigma_{\epsilon_{a_{k}}^{*}}^{2} \right)f\left( \delta_{jk,1}=c \right)}{\sum_{c^{'}=0}^{1} f\left( \hat{\mathbf{b}},{\hat{\mathbf{a}}}_{k},\boldsymbol{|}\delta_{jk,1}=c^{'},\mathbf{u}_{jk},\boldsymbol{\Sigma}_{jk},\sigma_{e_{\beta}^{*}}^{2},\sigma_{\epsilon_{a_{k}}^{*}}^{2} \right)f\left( \delta_{jk,1}=c^{'} \right)}, c=0,1\#(S32)\# \end{aligned}$$

Where $f\left( \hat{\mathbf{b}},{\hat{\mathbf{a}}}_{k}\boldsymbol{|}\delta_{jk,1}=c,\mathbf{u}_{jk}\boldsymbol{,}\boldsymbol{\Sigma}_{jk}\boldsymbol{,}\sigma_{e_{\beta}^{*}}^{2},\sigma_{\epsilon_{a_{k}}^{*}}^{2} \right)=\int f\left( \hat{\mathbf{b}},{\hat{\mathbf{a}}}_{k}\boldsymbol{|}\delta_{jk,1}=c,\mathbf{u}_{jk}\boldsymbol{,}\boldsymbol{\Sigma}_{jk}\boldsymbol{,}\sigma_{e_{\beta}^{*}}^{2},\sigma_{\epsilon_{a_{k}}^{*}}^{2} \right) f\left( u_{jk,1}|{\delta_{jk,1},u}_{jk,2} \right)du_{jk,1}$. When “1” denotes the complex trait, $f\left( \delta_{jk,1}=1 \right)=\pi_{\beta}$, and when “1” denotes the $k$-th molecular phenotype, $f\left( \delta_{jk,1}=1 \right)=\pi_{\alpha}$.

The full conditional distribution of $\boldsymbol{\Sigma}_{jk}$ in the summary-level model with a low-rank LD matrix is the same as that in the individual-level model.

The full conditional distribution of residual $\sigma_{e_{\beta}^{*}}^{2}$ is

$$\begin{aligned} f\left( \sigma_{e}^{2}|\hat{\mathbf{b}} \right)&\propto f\left( \hat{\mathbf{b}}|\sigma_{e}^{2} \right)f\left( \sigma_{e}^{2} \right) \\ &\propto\left( \sigma_{e}^{2} \right)^{-\frac{n_{g}}{2}} exp\left\{ -\frac{\left( \hat{\mathbf{b}}-\mathbf{Q}_{\mathbf{G}}\boldsymbol{\gamma}-\mathbf{Q}_{\mathbf{X}}\boldsymbol{\beta} \right)^{T}\left( \hat{\mathbf{b}}-\mathbf{Q}_{\mathbf{G}}\boldsymbol{\gamma}-\mathbf{Q}_{\mathbf{X}}\boldsymbol{\beta} \right)}{2\sigma_{e}^{2}} \right\}\left( \sigma_{e}^{2} \right)^{-\frac{\nu_{e}+2}{2}}exp\left\{ -\frac{\nu_{e}S_{e}^{2}}{2\sigma_{e}^{2}} \right\} \\ &\propto\left( \sigma_{e_{\beta}}^{2} \right)^{-\frac{n_{g}+\nu_{e}+2}{2}} exp\left\{ -\frac{\boldsymbol{SSE}_{e}+\nu_{e}S_{e}^{2}}{2\sigma_{e}^{2}} \right\} \\ &\sim\chi^{-2}\left( \hat{\nu}_{e},\hat{S}_{e}^{2} \right)\#(S33)\# \end{aligned}$$

Where $e=e_{\beta}^{*}$, $\tilde{\nu}_{e}=n_{g}+\nu_{e}$ and $\tilde{S}_{e}^{2}=\left( \boldsymbol{SSE}_{e}+\nu_{e}S_{e}^{2} \right)/{\tilde{\nu}_{e}}$.

The full conditional distribution of residual $\sigma_{\boldsymbol{\epsilon}_{k}}^{2}$ is

$$\begin{aligned} f\left( \sigma_{\boldsymbol{\epsilon}_{k}}^{2}|{\hat{\mathbf{a}}}_{k}, \right)&\propto f\left( {\hat{\mathbf{a}}}_{k}|\sigma_{\epsilon_{k}}^{2} \right)f\left( \sigma_{\epsilon_{k}}^{2} \right)\sim\chi^{-2}\left( \hat{\nu}_{e},\hat{S}_{e}^{2} \right)\#(S34) \end{aligned}$$

Where $e=\epsilon_{a_{k}}^{*}$, $\tilde{\nu}_{\boldsymbol{\epsilon}_{k}}=n_{k}+\nu_{\boldsymbol{\epsilon}_{k}}$, $\tilde{S}_{\boldsymbol{\epsilon}_{k}}^{2}=\left( \boldsymbol{SSE}_{\boldsymbol{\epsilon}_{k}}+\nu_{\boldsymbol{\epsilon}_{k}}S_{\boldsymbol{\epsilon}_{k}}^{2} \right)/{\tilde{\nu}_{\boldsymbol{\epsilon}_{k}}}$, and $\boldsymbol{SSE}_{\boldsymbol{\epsilon}_{k}}=n_{k}\left\{ \frac{\mathbf{y}^{T}\mathbf{y}}{n_{k}}\boldsymbol{-}\boldsymbol{\alpha}_{k}^{T}\left( \mathbf{a}_{k}\boldsymbol{+}\mathbf{Q}_{z}^{T}\left[ \boldsymbol{\Lambda}_{\mathbf{Z}}^{\boldsymbol{-}\frac{\boldsymbol{1}}{\boldsymbol{2}}}\mathbf{U}_{\mathbf{Z}}^{T}\mathbf{a}_{k}\boldsymbol{-}\mathbf{Q}_{\mathbf{Z}}\boldsymbol{\alpha}_{k} \right] \right) \right\}$.
